# A FinnGen pilot clinical recall study for Alzheimer’s disease

**DOI:** 10.1101/2023.02.06.23285534

**Authors:** Valtteri Julkunen, Claudia Schwarz, Juho Kalapudas, Merja Hallikainen, Aino-Kaisa Piironen, Arto Mannermaa, Hanna Kujala, Timo Laitinen, Veli-Matti Kosma, Teemu I. Paajanen, Reetta Kälviäinen, Mikko Hiltunen, Sanna-Kaisa Herukka, Sari Kärkkäinen, Tarja Kokkola, Mia Urjansson, Finn Gen, Markus Perola, Aarno Palotie, Eero Vuoksimaa, Heiko Runz

**Author notes:** equal contribution (Eero Vuoksimaa, Heiko Runz).

## Abstract

**Background:** Successful development of novel therapies requires that clinical trials are conducted in patient cohorts with the highest benefit-to-risk ratio. Population-based biobanks with comprehensive health and genetic data from large numbers of individuals hold promise to facilitate identification of trial participants, particularly when interventions need to start while symptoms are still mild, such as for Alzheimer’s disease (AD). However, few studies have yet tested whether recalling biobank participants into clinical follow-up studies is feasible.

**Objective:** To establish a process for clinical recall studies from FinnGen and demonstrate the feasibility to systematically ascertain customized clinical data from FinnGen participants with ICD10 diagnosis of AD or mild cognitive disorder (MCD).

**Methods:** Single-center cross-sectional study testing blood-based biomarkers and cognitive functioning in-person, computer-based and remote.

**Results:** 19% (27/140) of a pre-specified FinnGen subcohort were successfully recalled and completed the study. Hospital records largely validated registry entries. For 8/12 MCD patients, other reasons than AD were identified as underlying diagnosis. Cognitive measures correlated across platforms, with highest consistencies for dementia screening (r=0.818) and semantic fluency (r=0.764), respectively, for in-person versus telephone-administered tests. GFAP (p<0.002) and pTau-181 (p<0.020) most reliably differentiated AD from MCD participants.

**Conclusions:** Informative, customized clinical recall studies from FinnGen are feasible.

## INTRODUCTION

Alzheimer’s disease (AD) is the most common neurodegenerative condition and a major socio-economic challenge. It has been estimated that during the next four decades the prevalence of AD will quadruple from 27 to 106 million by which time 1 in 85 individuals worldwide will be living with the disease. Even a modest delay of disease onset could reduce the number of cases substantially (1). Interventions for AD are expected to be most efficacious when initiated in prospective patients at the earliest stages of disease (2). However, due to the lack of curative therapies or easily measurable biomarkers, it is current clinical practice that a majority of the patients receives their diagnosis only at progressed stages when irreversible alterations to brain and cognitive performance have already occurred. There is thus a substantial interest across industry and academia to identify AD patients early, and to align on measures that reliably diagnose the disease at the earliest possible stages to refine its onset and predict its course and response to experimental treatments.

Population-based, large-scale biobank studies that link detailed longitudinal health information with genetic and biomarker data are expediting the development of tools to identify individuals with a high probability to develop AD and other late onset diseases (3–5). They further hold the promise to accelerate recruitment of newly diagnosed patients or individuals at risk into observational and interventional clinical trials, or to facilitate access to efficient treatments (6,7). However, thus far few studies have overcome the scientific, logistical, and ethical challenges to recall individuals consented for broad population-level biobank research into customized clinical follow-up studies based on their individual-level health, genetic, or biomarker data.

The FinnGen (FG) study (www.finngen.fi) is a precompetitive partnership between thirteen industry partners and the Finnish biobanks to genetically profile and link genetics with detailed biomedical and social information in 10% of the Finnish population. Since its inception in 2017, FG has recruited over 500,000 participants with registry data on more than 2,000 harmonized disease endpoints and contributed to numerous scientific discoveries (8,9). FG participants are broadly consented for secondary use of research data and in principle re-contactable for follow-up studies through participating biobanks.

Here, we established a process for recalling FG participants fulfilling distinct, pre-specified inclusion criteria into a clinical follow-up study to obtain customized novel clinical and biomarker data. In a single-center, cross-sectional pilot study, we targeted a population that based on registry entries had been diagnosed (ICD10) with either AD or mild cognitive disorder (MCD). We assessed the accuracy of FG registry versus hospital records, applied three different approaches to evaluate cognitive functioning via in-person neuropsychological assessment, computerized and telephone-administered testing, and obtained blood-based AD biomarkers. Our results validate the accuracy of clinical information captured in Finnish registries, support remote cognitive testing as an effective screening approach, and demonstrate that recall studies from FG are feasible.

## MATERIALS AND METHODS

### Study participant selection

For this pilot study we targeted a FG subcohort with registry-defined diagnosis of either a wide definition of AD (ICD10: G30), or a registry entry of the ICD10 code F06.7 “Mild neurocognitive disorder due to known physiological condition”, the closest diagnostic code to mild cognitive impairment, a heterogenous condition frequently used to describe individuals with a higher probability to develop AD. Moreover, study participants were required to have provided consent for recontact, as well as for sample and data usage to the Biobank of Eastern Finland (BEF). The process for accessing biobank data and steps required to operationalize a biobank recall study is guided by the Finnish biobank regulations (https://ita-suomenbiopankki.fi/en/researchers/) and visualized in **Figure 2**. In brief, after a positive feasibility assessment by BEF, approval of the study protocol and accompanying documents by the ethics committee of Helsinki University Hospital, and ensuring a permit to access patient files from Kuopio University Hospital (KUH), a request for biobank data access was launched via the Fingenious portal (https://site.fingenious.fi/). This triggered a formal evaluation of the study plans by the BEF scientific steering committee and initiation of contracts between participating parties. An updated feasibility assessment by BEF resulted in 150 participants meeting study inclusion criteria. Of these, eight were removed as they were identified as duplicates (same person counted twice in the registry). Two additional individuals were excluded since they lived in institutionalized care with a likely late-stage AD and not being able to travel to the study site. The study participant selection with exclusions is presented in **Figure 1a**.

**Figure 1.**
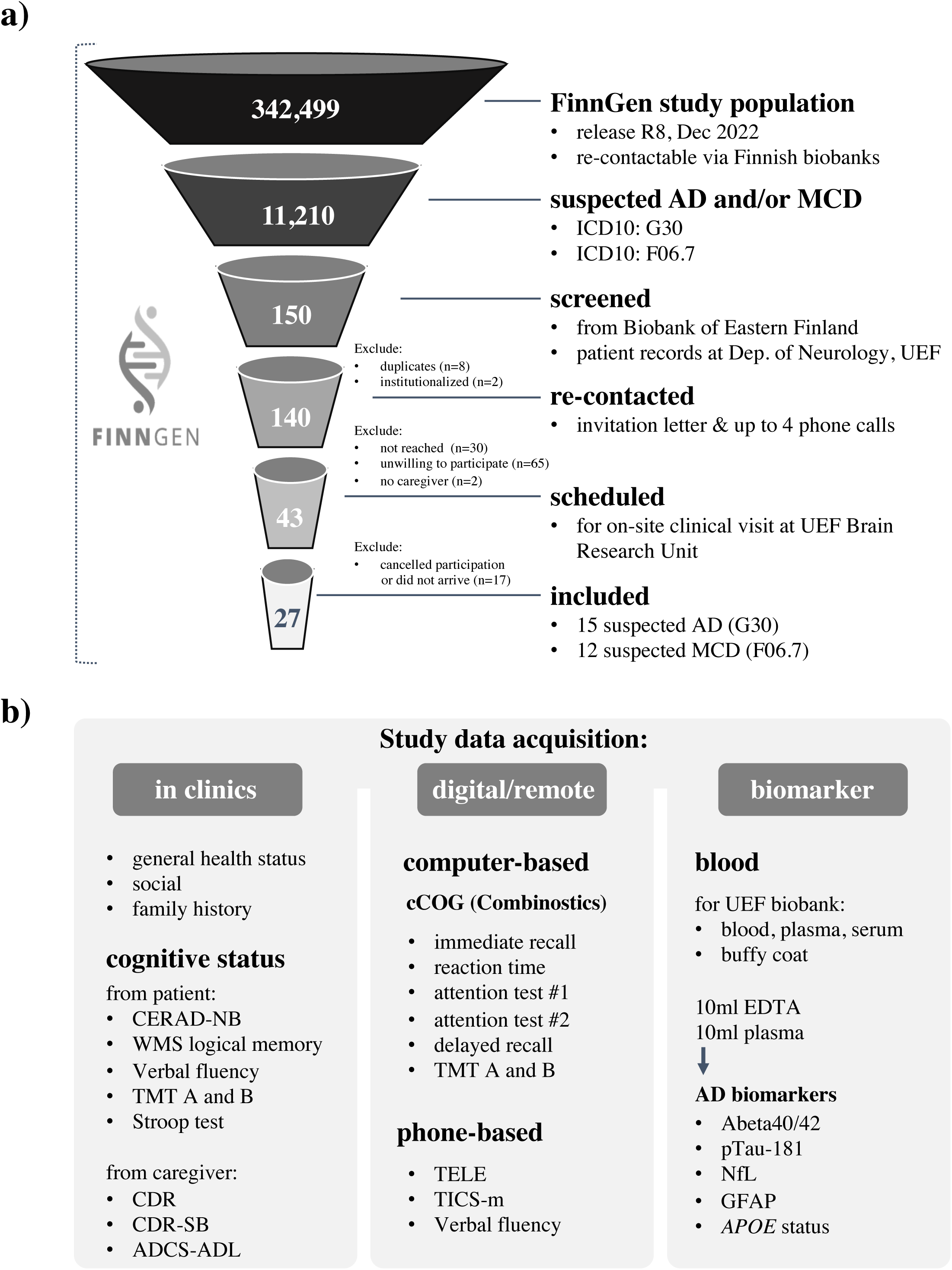
a) Screening and enrollment process for study. AD, Alzheimer’s disease; MCD, mild-cognitive disorder; UEF, University of eastern Finland. **b)** New data acquired during study. CERAD-NB, Consortium to Establish a Registry for Alzheimer’s Disease Neuropsychological Battery; WMS, Wechsler Logical Memory Scale; TMT-A/B, Trail Making Test parts A and B; CDR, clinical dementia rating scale; CDR-SB, CDR sum of boxes; ADCS-ADL, Alzheimer’s Disease Cooperative Study – Activities of Daily Living scale; TELE, telephone assessment for dementia; TICS-m, modified Telephone Interview for Cognitive Status; Abeta-42/40, amyloid beta (Aβ)1-42/1-40; p-Tau181, phosphorylated-tau181; NfL, neurofilament light chain; GFAP, glial fibrillary acidic protein;

**Figure 2.**
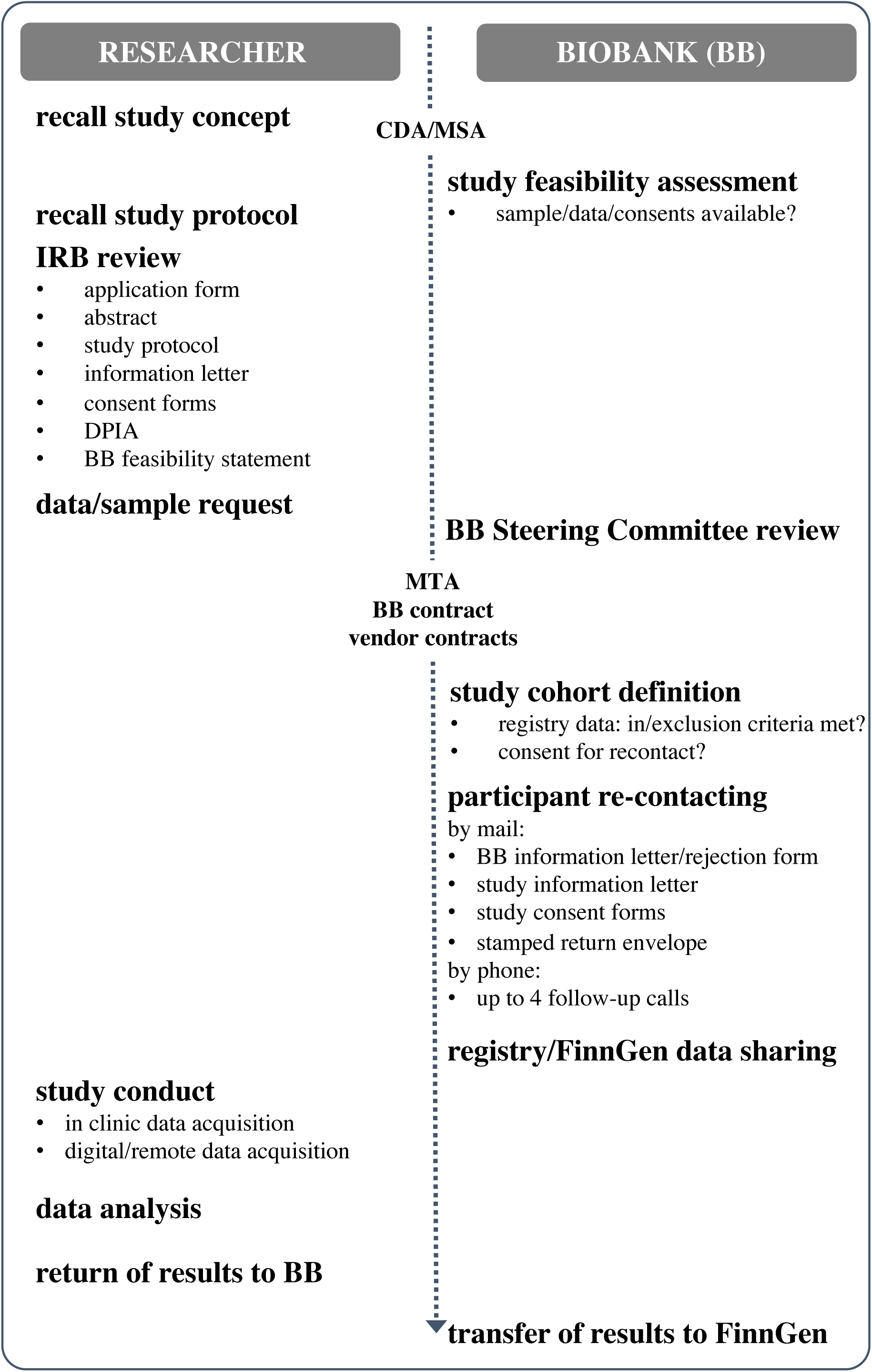
Operationalization of this clinical recall study from FinnGen. CDA, confidentiality agreement; MSA, master services agreement; IRB, institutional review board; DPIA, data protection impact assessment; BB, biobank; MTA, material transfer agreement

A total of 140 FG participants were contacted by BEF personnel. Recontact was conducted by using a letter that included study information and forms for informed written consent. To ensure that the study information had reached the potential study participants, and to further enhance the participation rate, all potential participants received a phone call from the BEF study nurse within 1-2 weeks after sending the invitation material. During this phone call the potential participants received standard structured information concerning the study, were asked whether they were willing to participate, and a date for on-site assessment at the University of Eastern Finland (UEF) Brain Research Unit (BRU) was scheduled. Participants willing to participate returned their informed written consents either by mail to BRU, or by bringing the signed forms to their appointment. Participants who completed the in-person and computerized cognitive testing batteries at the BRU received a follow-up phone-call after 1-2 weeks with the aim to perform a remote telephone-administered cognitive assessment. Data obtained during this study are summarized in **Figure 1b**.

### Schedule of assessment

All study participants were invited to UEF-BRU for: a structured interview of the study participant and caregiver or family member to obtaining basic patient information and to fill out questionnaires regarding abilities of daily life and functioning; drawing of blood samples for AD biomarker profiling and optional future biomarker discovery; in-person neuropsychological assessment performed by a trained psychologist; and on-site computer-administered cognitive function testing. Moreover, cognitive function was subsequently evaluated by phone-interview 1-2 weeks later. All cognitive assessments were done in Finnish language. Half of the study participants were randomized to obtain computerized-testing before in-person testing to reduce the risk of systematic bias from practice effect.

### Data linkage to FinnGen and hospital records

In addition to obtaining new clinical and biomarker data, we obtained approval to link these data to previously ascertained health care register data such as hospital diagnoses, medication records, social registry entries, and genetic data. Data linkage was performed in a controlled FG Sandbox environment approved to host individual-level data and accessible only by approved researchers. In addition to data directly ascertained through the study protocol or available through FG, we also analyzed earlier routine care data available for study participants through the patient records from KUH. This data included clinical information from earlier memory clinic and follow-up visits (e.g., historic cognition test results), previous laboratory values (e.g., results from cerebrospinal fluid examinations for AD biomarkers), and neuroimaging data (e.g., magnetic resonance imaging ascertained through routine care). Among others, data linkage offered the possibility to compare the accuracy of FG registry data against hospital records and validate that inclusion criteria were indeed met based on earlier clinical information.

### In-person neuropsychological testing

In-person neuropsychological testing included the Consortium to Establish a Registry for Alzheimer’s Disease Neuropsychological Battery (CERAD-NB) (10). Additional tests included Logical Memory subtest from the Wechsler Adult Intelligence Scale 3^rd^ edition (11), Trail Making Test A (TMT-A) and B (TMT-B) (12), and a short 40-item version of the Stroop test (13). The tests were administered by a trained psychologist at the UEF-BRU. For the current study we investigated the following four specific cognitive domains (with specific measures in parentheses): episodic memory (immediate free recall of words in trials 1-3 and delayed free recall of the 10-word list); executive function (set-shifting: TMT-B controlled for TMT-A); processing speed (TMT-A); and semantic fluency (1-minute animal naming). These measures were selected because similar or corresponding measures were also available in computerized testing and in the telephone interview (described below).

### Computerized cognitive testing

We used the cCOG (14) platform developed by Combinostics Oy (https://www.cneuro.com), a self-administered computer-based testing tool for assessing cognitive performance at an on-site computer via a keyboard and a mouse. A total of seven tasks measured performance in four specific cognitive domains: episodic memory, executive function, processing speed, and reaction time. An earlier study proposed that cCOG has comparable accuracy to in-person neuropsychological batteries for detecting dementia, and at least moderately correlates with specific in-person neuropsychological tests (14). In this current study, we used episodic memory measures of immediate (total words recalled in trials 1-3) and delayed recall of 12-word list measures. We also used the Modified TMT, with two parts corresponding to the traditional TMT-A and -B, to measure processing speed and executive function (set-shifting). Detailed test descriptions and protocols can be found in Rhodius-Meester et al. (2020)(14). Completing cCOG took participants about 25 minutes.

### Telephone interview to assess cognitive performance

We used two validated, telephone-administered cognitive screening instruments: telephone assessment for dementia (TELE) (15) and the modified Telephone Interview for Cognitive Status (TICS-m) (16). Telephone interviews took on average 17 (range 12-33) minutes with an average of 13 (range 9-29) minutes to complete TELE and TICS-m and were conducted by a trained research nurse. For analyses in the current study, we used the following measures: For TELE, we used a continuous score (0-20) and validated cut-offs for cognitive impairment (<16) and healthy cognition (>17.5) (17). From TICS-m, we used episodic memory measures of verbal immediate (total words recalled in trials 1-3) and delayed free recall of 10-word list. In the original TICS-m, there is only a single trial of the 10-word list in the learning condition. Here we modified this task to include three learning trials in order to make this measure more similar to in-person and computerized list learning tasks. In addition, semantic fluency was measured (animals in 1-minute).

### Assessment of functional status and independence in activities of daily living

The severity of cognitive symptoms and functioning disability was estimated by the clinical dementia rating scale (CDR) and CDR sum of boxes (CDR-SB) (18) as well as by the 18-item Alzheimer’s Disease Cooperative Study – Activities of Daily Living scale (ADCS-ADL) (19). This on-site assessment was done by a trained study nurse and included an interview of the caregiver or family member of the study participant to increase the reliability of data acquisition.

### AD blood-based biomarkers and *Apolipoprotein E (APOE)* genotype

The analyses of plasma biomarkers phosphorylated-tau181 (pTau-181), amyloid beta (Aβ)1-40, Aβ1-42, glial fibrillary acidic protein (GFAP) and neurofilament light chain (NfL) from all 27 patients were performed using Simoa HD-X analyzer (Quanterix, Billerica, Massachusetts, USA), which employs single molecule array digital immunoassay technology. Analyses were performed at the biomarker laboratory of UEF-BRU. Plasma pTau-181 levels were quantified using Simoa pTau-181 Advantage V2 Kit (Ref# 103714, Quanterix) and Aβ1-40, Aβ1-42, GFAP and NfL levels using Simoa Neurology 4-Plex E Advantage Kit (Ref# 103670, Quanterix) according to the manufacturer’s instructions. Prior to analyses, EDTA plasma samples were thawed, mixed, centrifuged (10,000xg, 5 min, +20°C) and measured in duplicates. All replicates had coefficient of variation below 15%. *APOE* status was defined by two single-nucleotide polymorphisms, rs429358 and rs7412, in chromosome 19. We classified individuals as ε4-carriers vs. non-carriers.

### Statistical analyses

We compared AD and MCD groups with regards to cognitive function and performed comparisons between in-person, computerized and telephone-administered tests. Cognitive screening instruments were the Mini Mental State Examination (MMSE) as a part of CERAD-NB battery administered in-person and TELE included in the telephone interview. We also classified individuals in cognitively impaired versus cognitively intact based on the CERAD-NB word list learning immediate and delayed recall measures. With regards to specific abilities, we compared performance in the following cognitive domains (measured on at least two platforms): episodic memory, executive function (set-shifting), processing speed, and semantic fluency. Episodic memory was measured in all three formats (CERAD word list in in-person, TICS 10-word list via telephone and 12-word list in cCOG), processing speed and executive function in two formats (TMT-A & TMT-B in in-person and modified TMT-A & TMT-B in cCOG), and semantic fluency in two formats (1-minute animal fluency both in in-person and telephone testing). Statistical analyses were performed with R version 4.1.1 (available online at https://www.R-project.org/). Spearman’s rank partial correlation analyses were used to test the associations between in-person, computerized and telephone-administered tests. We report Spearman correlations due to small sample size and non-normal distribution of some of the measures. In addition, cognitive test results, age, and the time from original diagnosis to study visit were compared between AD and MCD groups using independent t-tests. The significance level was set at α = 0.05.

### Ethics approvals

All participants in this recall study were also participants in FG and had provided informed consent for biobank research including an option for recalling into a new scientific study. This procedure is based on the Finnish Biobank Act. All recall study participants and their respective caregiver or family member received detailed information regarding this study by a formal information letter, as well as structured information provided during a phone call. All study participants and their participating caregivers were asked for and provided written informed consent. All study data was initially stored locally at UEF (on-site assessment) and the Finnish Institute for Molecular Medicine (FIMM) (remote cognition assessment by phone). Study results and data were then returned first to BEF and from there further to the FG Sandbox controlled environment to link newly ascertained clinical data with genetic and register data. The recall study was reviewed and approved by the ethics committee of HUS under IRB approval number 990/2017. Additionally, research permissions were granted from BEF (diary ID 1248/2021) and KUH (1535/2021, study ID 5772671).

An alternative process for dedicated biobank consent has been applied for a subset of FG participants. Some of the separate legacy research cohorts collected prior the Finnish Biobank Act came into effect (in September 2013) and start of FG (August 2017) were collected based on previous study-specific consents and later transferred to the Finnish biobanks and eventually to FG after approval by Fimea (Finnish Medicines Agency) and the National Supervisory Authority for Welfare and Health. Recruitment protocols followed the biobank protocols approved by Fimea. The Coordinating Ethics Committee of the Hospital District of Helsinki and Uusimaa (HUS) statement number for the FinnGen study is Nr HUS/990/2017. The permit numbers of the decisions made by Finnish Institute for Health and Welfare and the Biobank Access Decisions for FG samples and data utilized in FG Data Freeze 9 are presented in Acknowledgements.

## RESULTS

### Operationalization of a FinnGen clinical recall study

At the time of study (October 2021-February 2022), FG had ascertained genetic and registry data on 392,499 participants (190,879 females, 151,620 males; DF8, publicly released in December 2022 via http://finngen.fi/en/accesss_results). Of these, health records from 11,210 participants were characterized by at least one entry of the FG diagnostic code “G6_ALZHEIMER” (curated based on ICD10 code G30), reflecting a putative diagnosis of AD. 2,170 participants had at least one entry for ICD10 code F06.7, reflecting a diagnosis of MCD (http://risteys.finngen.fi/). For this proof-of-concept recall study, we decided to focus on participants recruited into FG from just one of the nine contributing biobanks, Biobank of Eastern Finland (BEF; (http://ita-suoimenbiopankki/en/). To enable comparison of FG registry data with individual-level health records and limit geographic outreach, we further requested that study participants were still alive, lived within a proximity of 100km from the study site and had earlier been a patient at the Department of Neurology, KUH.

A flow-chart visualizing study participant selection and data ascertained during study are provided in **Figure 1**, and the process how this FG recall study was operationalized is described in **Figure 2** and Methods. In brief, over a period of 15 business days, 277 phone calls were made (maximum of 3-4 calls/person) of which 151 were not answered. A total of 65 individuals were reached and declined to participate, 30 individuals were not reached, and for two individuals no caregiver could be identified to accompany the in-principle-interested participant to a near-term on-site visit. This resulted in a total of 43 FG participants (40% of the 108 reached and eligible) for whom a study visit at the UEF-BRU was scheduled. A total of 27 on-site visits were performed in November and December 2021. The initial plan to continue recruitment of this particularly vulnerable population until the end of February 2022 was discontinued after the majority of scheduled visits were canceled at the peak of a SARS-Cov2 outbreak in Finland in early 2022. With this, 63% (27/43) of the scheduled participants came to study visit and the participation rate of this first FG clinical recall study was 19% (27/140) when considering the entire *a-priori* eligible and re-contactable cohort, but it likely would have been substantially higher if all scheduled study visits could have been completed in 2021.

### Registry diagnoses and hospital records are largely consistent

Of the 27 FG participants included in the study, 13 were females (**Table 1**). The mean age at study visit was 73 (58-87) years. Based on FG registry data, 15 study participants had a diagnosis of AD, while 12 participants had been diagnosed with MCD. Mean age at the first diagnostic entry of either AD or MCD was 69 (50-81) and 69 (58-80), respectively, with the mean time since diagnosis being 4 (1-13) years for AD and 3 (1-6) years for the MCD patients under study. Based on these measures, our subcohort was largely representative of the broader FG AD and MCD populations (http://risteys.finngen.fi/).

**Table 1.** Characteristics of the study cohort.

|  | AD (n=15) | MCD (n=12) | p-value |
| --- | --- | --- | --- |
| Age, y | 73.0<br>(8.8) | 72.5 (8.3) | 0.895 |
| Sex, females/males [n] | 8/7 | 5/7 | 0.830 |
| APOE4 carrier [n] | 10 | 3 | 0.171 |
| MMSE | 24.0 (7.6) | 24.1 (6.7) | 0.977 |
| In-person tests |  |  |  |
| CERAD immediate recall | 14.6 (6.6) | 15.2 (6.0) | 0.812 |
| CERAD delayed recall | 4.0 (2.8) | 5.0 (2.4) | 0.343 |
| TMT-A [s] | 84.3 (54.2) | 83.6 (35.0) | 0.973 |
| TMT-B [s] | 178.1 (83.9) | 215.2 (83.8) | 0.337 |
| Semantic fluency | 18.1 (7.3) | 18.0 (8.2) | 0.963 |
| Computer-based tests |  |  |  |
| cCOG immediate recall | 13.6 (6.2) | 12.6 (5.1) | 0.666 |
| cCOG delayed recall | 3.5 (3.2) | 4.1 (2.8) | 0.632 |
| cCOG TMT-A modified [s] | 98.8 (74.1) | 81.1 (33.6) | 0.567 |
| cCOG TMT-B modified [s] | 231.6 (87.4) | 261.0 (91.6) | 0.525 |
| Phone-based tests |  |  |  |
| TELE total | 15.3 (4.9) | 17.2 (4.9) | 0.346 |
| TICS-m immediate recall | 14.5 (9.6) | 14.2 (6.6) | 0.927 |
| TICS-m delayed recall | 3.8 (4.0) | 2.6 (2.4) | 0.365 |
| Semantic fluency | 15.1 (6.8) | 17.2 (6.7) | 0.446 |

Data are given as mean and standard deviation for Alzheimer's disease (AD) and mild cognitive disorder (MCD) groups separately. *APOE4* carrier status was available for 23 (14AD/9MCD) and MMSE, CERAD immediate and delayed recall, cCOG immediate and delayed recall, and in-person verbal fluency data was available for 26 (14AD/12MCD) participants. In-person TMT-A was available for 25 (14AD/11MCD), in-person TMT-B for 20 (11AD/9MCD), cCOG TMT-A for 17 (10AD/7MCD), and cCOG TMT-B for 17 (11AD/6MCD) participants. APOE, Apolipoprotein E, cCOG, computerized cognitive testing; CERAD, Consortium to Establish a Registry for Alzheimer's Disease; MMSE, Mini Mental State Examination; TELE, Telephone assessment for dementia; TICS-m, modified Telephone Interview for Cognitive Status with three learning trials of the 10-word list; TMT-A/B, Trail Making Test parts A and B.

For all participants, the dates of diagnosis between FG registry entry and hospital records were within a six-month range. For one participant, F06.7 was only documented in registry, but not in hospital records. According to hospital memory clinic records, only a single patient had progressed to AD from an initial diagnosis of MCD. For the majority of the MCD group, clinical interpretation of the hospital records could pinpoint at least one alternative etiology explaining the mild cognitive impairment. This included one case with vascular cognitive impairment, one with cortico-basal degeneration and one with idiopathic normal pressure hydrocephalus. Other putative reasons for MCD included Parkinson’s disease, depression, epilepsy, menopause, sleeping problems, or tiredness due to poor overall health. Conversely, for 13 of the 15 patients in the AD group, the clinical diagnosis had been based on current Finnish diagnostic guidelines for memory disorders (20), which included administration of a formal neuropsychological testing battery (CERAD-NB or comprehensive neuropsychological assessment), brain imaging with magnetic resonance imaging or computed tomography, thorough neurological and physical examination as well as in several cases cerebrospinal fluid biomarkers and brain positron emission tomography. Two participants in the AD group did not have diagnostic information in hospital records. For these cases, the validity of a prior diagnosis of AD could not be confirmed. Three had documentation for mixed AD: in one case AD presented together with vascular dementia features, in another AD was documented together with Lewy-Body dementia, and one case was documented to have a posterior variant of AD.

### In-person, computerized and telephone-administered assessment of cognition

Data acquisition via all three platforms was in general feasible for participants, with no substantial differences between the AD and MCD groups, although several individuals from both groups were unable to complete specific subtests. Most problems occurred with the computerized versions of TMT-A and TMT-B which were completed by only 17 participants.

In-person MMSE scores could be obtained from 26 participants. Of these, 17 (8AD/9MCD) showed normal cognitive function or only mild impairment (MMSE>=25), while 9 (6AD/3MCD) were classified as cognitively impaired (MMSE<25). Although the mean MMSE scores did not differentiate between AD (24.0±7.6) and MCD (24.1±6.7) groups (p=0.977, **Table 1**), there were more cognitively impaired individuals in the AD group (43%, 6/14 versus 25%, 3/12 for MCD).

According to TELE, 16 participants (8AD/8MCD) were classified as cognitively healthy (cut-off score >17.5), while 7 (5AD/2MCD) participants were classified as cognitively impaired (cut-off score <16), and 4 (2AD/2MCD) participants ranked between the cut-off scores (<17.6 and >=16). On average those with AD (15.3±4.9) had non-significantly (p=0.345) lower scores than those with MCD (17.2±4.9). The prevalence of cognitively impaired was almost double in the AD group (33%, 5/15) than in the MCD group (17%, 2/12).

Based on CERAD-NB word list immediate recall, 71% (10/14) of those with AD and 50% (6/12) of those with MCD were classified as having episodic memory impairment (cut-off <17), with mean immediate recall scores of 14.6±6.6 and 15.2±6.0 in AD and MCD subgroups, respectively (p=0.812). Based on the CERAD-NB word list delayed recall, 57% (8/14) of those with AD and 33% (4/12) of those with MCD were classified as having episodic memory impairment (cut-off <5), with mean scores of 4.0±2.8 and 5.0±2.4 in AD and MCI subgroups, respectively (p=0.343).

Continuous MMSE and TELE scores showed a high correlation of r=0.818 (p<0.001) (**Figure 3a**, **Table 1**). A similar high correlation was observed for semantic fluency as measured by in-person testing as well as a part of the telephone interview (r=0.764; p<0.001; **Figure 3b**). Considering measures of episodic memory, immediate word recall measures were positively correlated between all three platforms (all r’s≥0.584; p’s≤0.002). Correlations between delayed word recall measures were smaller, with CERAD-NB showing modest correlations with cCOG (r=0.390, p=0.049) and TICS-m (r=0.424, p=0.031) whereas the correlation between cCOG and TICS-m measures was small (r=0.250, p=0.218) (**Figure 3c**). Strong correlations were observed between in-person and computerized measures of executive function (TMT-B; r=0.689, p=0.004, n=15; r=0.536, p=0.089, n=11 after adjusting for TMT-A and r=0.62, p=0.054, n=10 after additional exclusion of TMT-A outliers), whereas in-person versus computerized measures of processing speed correlation was moderate (TMT-A; r=0.360, p=0.171, n=16) (**Figure 3d**).

**Figure 3.**
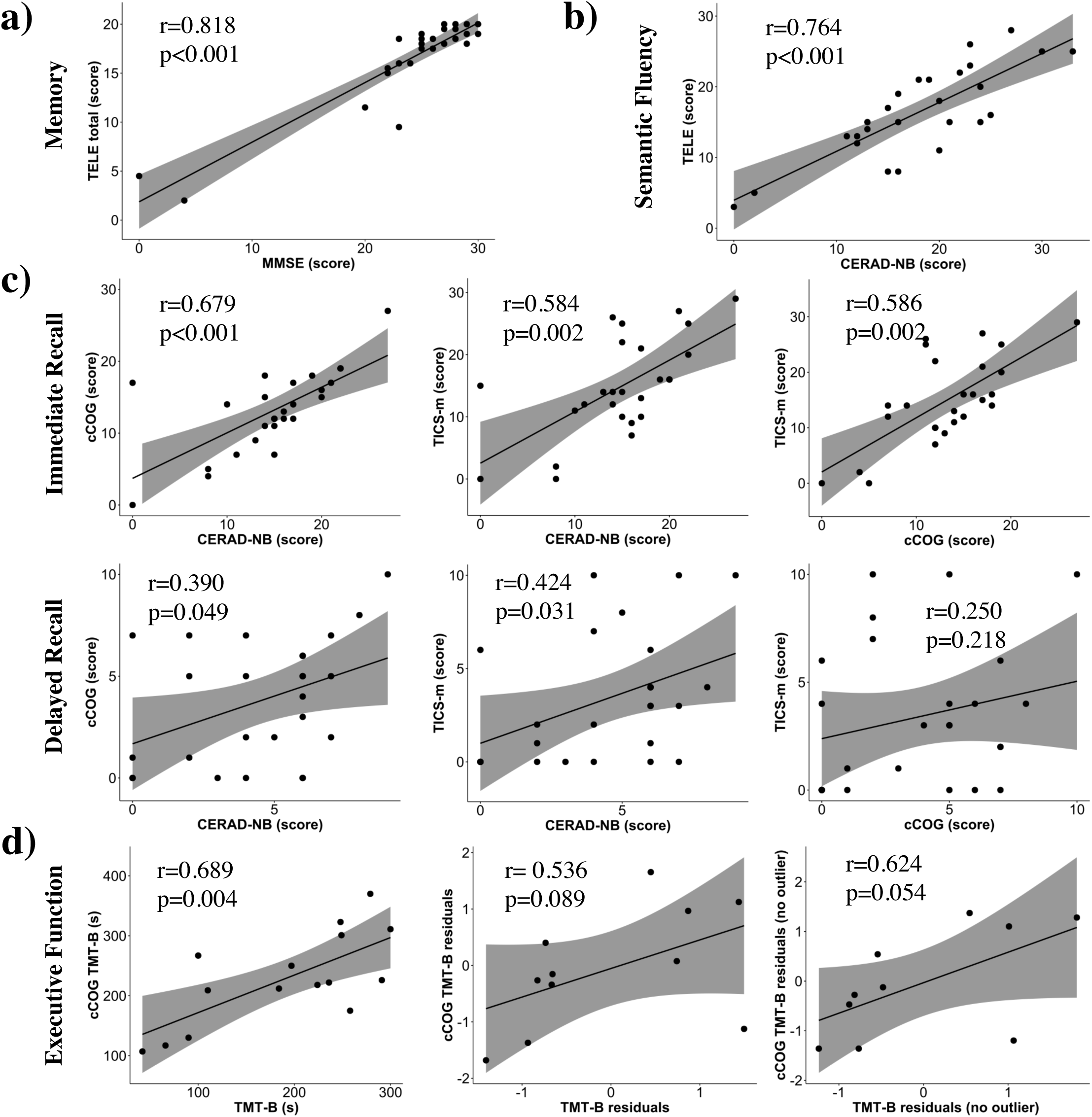
Comparison of cognitive testing results across the three platforms utilized in the study. Correlations of **a)** memory, **b)** semantic fluency, **c)** immediate and delayed recall, and **d)** executive function are presented between in-person, computerized, and telephone-based assessments. Scatterplots with 95% confidence intervals (shaded areas) display correlation coefficient r and p-value from Spearman’s rank partial correlation analyses. cCOG, computerized cognitive testing; CERAD-NB, Consortium to Establish a Registry for Alzheimer’s Disease Neuropsychological Battery; MMSE, Mini Mental State Examination; TELE, Telephone assessment for dementia; TICS-m, modified Telephone Interview for Cognitive Status with three learning trials of the 10-word list; TMT-A/B, Trail Making Test parts A and B.

### Blood-based biomarkers and *APOE* genotype

Across all blood-based biomarker parameters, values varied substantially (**Table 2**). Notably, despite the small cohort size we observed statistically significant differences between the AD and MCD groups for GFAP [p=0.002; AD: 216.3 (57.5-433.7) pg/ml versus MCD: 102.8 (30.5-161.3) pg/ml] and pTau-181 [p=0.020; AD: 3.8 (1.2-11.7) pg/ml versus MCD: 1.9 (0.6-3.6) pg/ml], whereas no significant differences were seen for Aβ42/40-ratio (p=0.266) or NfL (p=0.140). TELE total score was significantly correlated with GFAP (p=0.011), NfL (p=0.001) and pTau-181 (p=0.006), but not with Aβ42/40 (p=0.345), whereas non-significant correlations of MMSE score with biomarkers ranged from -0.247 to 0.020. With the exception of correlations between pTau-181 levels and telephone-administered immediate recall of the word list (r=-0.394, p=0.042, n=27) and in-person administered TMT-A (r=0.435, p=0.034, n=24) no other correlations between biomarkers and cognitive performance measures reached statistical significance threshold.

**Table 2.**
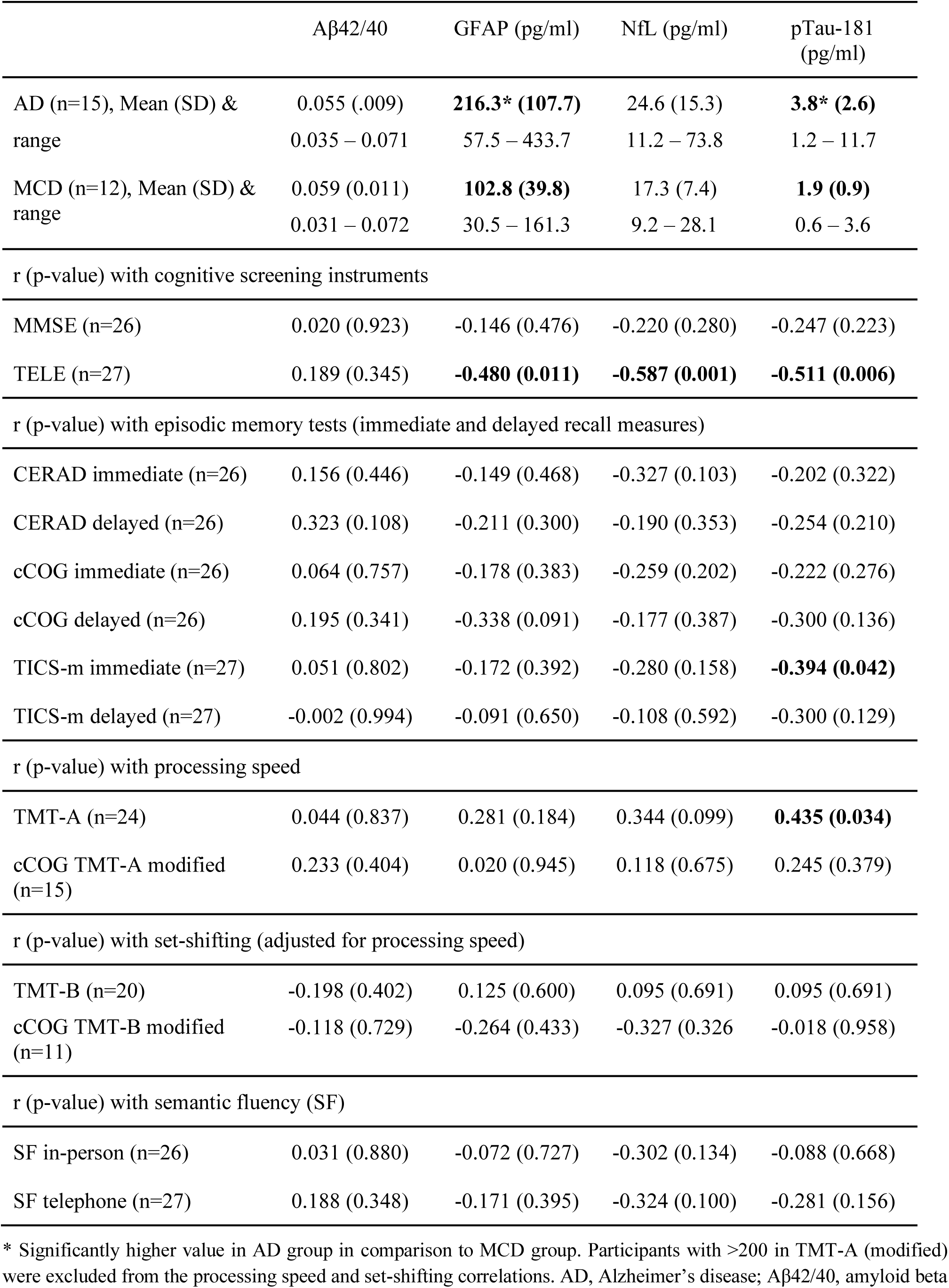
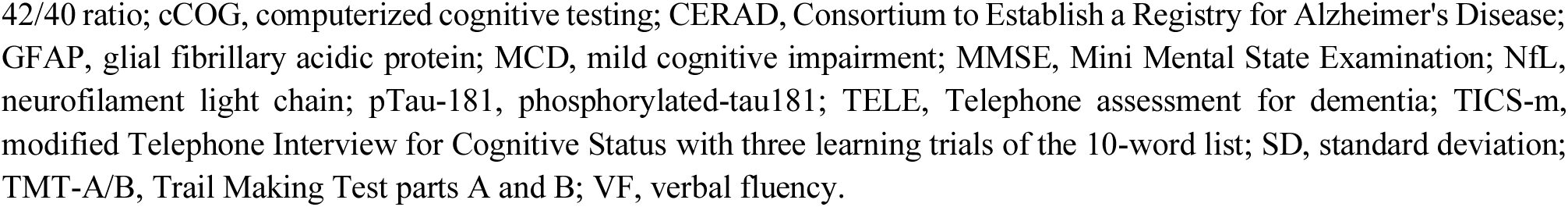
Blood-based biomarkers in study participants with Alzheimer’s disease (AD) and mild cognitive disorder (MCD) diagnosis and correlations with cognitive measures.

| | Aβ42/40 | GFAP (pg/ml) | NfL (pg/ml) | pTau-181 (pg/ml) |
| --- | --- | --- | --- | --- |
| AD (n=15), Mean (SD) & range | 0.055 (.009)<br>0.035 – 0.071 | **216.3\* (107.7)**<br>57.5 – 433.7 | 24.6 (15.3)<br>11.2 – 73.8 | **3.8\* (2.6)**<br>1.2 – 11.7 |
| MCD (n=12), Mean (SD) & range | 0.059 (0.011)<br>0.031 – 0.072 | **102.8 (39.8)**<br>30.5 – 161.3 | 17.3 (7.4)<br>9.2 – 28.1 | **1.9 (0.9)**<br>0.6 – 3.6 |
| r (p-value) with cognitive screening instruments |  |  |  |  |
| MMSE (n=26) | 0.020 (0.923) | -0.146 (0.476) | -0.220 (0.280) | -0.247 (0.223) |
| TELE (n=27) | 0.189 (0.345) | **-0.480 (0.011)** | **-0.587 (0.001)** | **-0.511 (0.006)** |
| r (p-value) with episodic memory tests (immediate and delayed recall measures) |  |  |  |  |
| CERAD immediate (n=26) | 0.156 (0.446) | -0.149 (0.468) | -0.327 (0.103) | -0.202 (0.322) |
| CERAD delayed (n=26) | 0.323 (0.108) | -0.211 (0.300) | -0.190 (0.353) | -0.254 (0.210) |
| cCOG immediate (n=26) | 0.064 (0.757) | -0.178 (0.383) | -0.259 (0.202) | -0.222 (0.276) |
| cCOG delayed (n=26) | 0.195 (0.341) | -0.338 (0.091) | -0.177 (0.387) | -0.300 (0.136) |
| TICS-m immediate (n=27) | 0.051 (0.802) | -0.172 (0.392) | -0.280 (0.158) | **-0.394 (0.042)** |
| TICS-m delayed (n=27) | -0.002 (0.994) | -0.091 (0.650) | -0.108 (0.592) | -0.300 (0.129) |
| r (p-value) with processing speed |  |  |  |  |
| TMT-A (n=24) | 0.044 (0.837) | 0.281 (0.184) | 0.344 (0.099) | **0.435 (0.034)** |
| cCOG TMT-A modified (n=15) | 0.233 (0.404) | 0.020 (0.945) | 0.118 (0.675) | 0.245 (0.379) |
| r (p-value) with set-shifting (adjusted for processing speed) |  |  |  |  |
| TMT-B (n=20) | -0.198 (0.402) | 0.125 (0.600) | 0.095 (0.691) | 0.095 (0.691) |
| cCOG TMT-B modified (n=11) | -0.118 (0.729) | -0.264 (0.433) | -0.327 (0.326) | -0.018 (0.958) |
| r (p-value) with semantic fluency (SF) |  |  |  |  |
| SF in-person (n=26) | 0.031 (0.880) | -0.072 (0.727) | -0.302 (0.134) | -0.088 (0.668) |
| SF telephone (n=27) | 0.188 (0.348) | -0.171 (0.395) | -0.324 (0.100) | -0.281 (0.156) |

\* Significantly higher value in AD group in comparison to MCD group. Participants with >200 in TMT-A (modified) were excluded from the processing speed and set-shifting correlations. AD, Alzheimer's disease; Aβ42/40, amyloid beta
42/40 ratio; cCOG, computerized cognitive testing; CERAD, Consortium to Establish a Registry for Alzheimer's Disease; GFAP, glial fibrillary acidic protein; MCD, mild cognitive impairment; MMSE, Mini Mental State Examination; NfL, neurofilament light chain; pTau-181, phosphorylated-tau181; TELE, Telephone assessment for dementia; TICS-m, modified Telephone Interview for Cognitive Status with three learning trials of the 10-word list; SD, standard deviation; TMT-A/B, Trail Making Test parts A and B; VF, verbal fluency.

*APOE* status was available for 23 participants. Of these 13 were *APOE* ε4-carriers (12 heterozygotes, 1 homozygote). There were 71% (10/14) and 33% (3/9) *APOE* ε4-carriers (p=0.072) in the AD and MCD groups, respectively (p=0.072 for the difference).

## DISCUSSION

In this single-center, cross-sectional pilot study we ascertained clinical-grade cognitive function and biomarker data from a pre-specified FG subcohort in order to build a framework to operationalize future clinical recall studies from FG. We demonstrated through targeted re-contacting of a FG subcohort with a registry-based diagnosis of AD or MCD that in our recall setting cognitive testing can be performed through on-site clinical examination, a customized web-based tool, or remotely through phone interview, and that measures of episodic memory, executive function and verbal fluency correlate across all three platforms. We further showed a high level of overlap between FG registry data and hospital records and further validate the diagnosis AD versus MCD by determining blood-based AD biomarkers from freshly acquired blood samples.

Enabling recall of biobank participants for clinical follow-up studies is a new frontier for biobank research and considered as of substantial potential across academic and industry stakeholders. In fact, it has been suggested that the “bottom-up” targeted recruitment of individuals with distinct systematically-acquired biomolecular findings is likely to substantially accelerate the speed to validate the relevance of human genetic variation for disease (22), or to generate functional insights on genetic discoveries. A few studies have given a glimpse on the potential of this approach. For instance, an earlier study in Finland in relatives of rare-variant carriers identified through population-level sequencing could substantiate the role of *SLC30A8* in insulin secretion and a Finn-enriched loss-of-function variant in that gene as protective for diabetes (23). Other examples include recall studies in the Estonian Biobank or East London Genes and Health that guided medical interventions (24) or informed transition of an early-stage rare disease therapeutic program towards the clinics (25). Recall studies from population biobanks also hold considerable promise to improve clinical development of precision medicine drugs. Currently, a large fraction of interventional studies fail projected enrollment timelines, a challenge that could potentially be overcome through targeted outreach towards distinct, well-characterized subsets of population biobank participants with the highest projected probability to qualify for and benefit from inclusion into a clinical trial (26). Despite such promises, numerous scientific, operational, and ethical challenges exist to firmly integrate biobank recall studies into the mainstream research, some of which impacted also to our study.

The more than 11,000 FG participants with a probable diagnosis AD at start of our study represent about 8% of prevalent dementia and AD cases in Finland (20). As a research cohort, these individuals already contributed to improve the understanding of the genetic etiology of AD (27,28). Utilizing research data from this cohort for a clinical recall study required thorough collaboration between the study investigators and BEF over a period of nearly two years from first contact to study completion. Specifically, a process needed to be developed that could in principle serve as a template for any future sample or clinical recall study from FG and the Finnish biobanks. Also, in accordance with the Finnish legislation, potential privacy concerns were mitigated by conducting all analyses in a secure compute environment, establishing recontact only through eligible biobank personnel, informing FG participants about their right to opt-out of continued use of their personal data for secondary use in research, and returning all results generated as part of a recall study to the respective biobank and FG for long-term storage and eventual public release.

While the sample size of our pilot recall study is too small for extrapolations to the entire FG population, it still speaks to the strength of the Finnish biobank system that for 93% (140/150) of the consented and targeted FG participants contact details were available, that 78% (110/140) could be reached through mail and recruitment calls within a period of less than three weeks, and that 30% (43/140) of patients and caregivers were in principle willing to enroll in the study. With 19% of contacted participants completing the study protocol, the final participation rate was in a similar range as that of an earlier questionnaire study where we had approached FG participants to provide cognitive, behavioral and lifestyle information via an online platform (29), despite our current study targeting an aged and diseased population with a high vulnerability to a concomitantly ravaging SARS-Cov2 outbreak. It can be expected that future observational studies from FG will yield higher enrollment rates by targeting a younger and healthier study population over an extended period of time and by returning information that can lead to impactful health and lifestyle changes.

Registry data proved to be highly consistent with hospital records in our study population, indicating that clinical data captured in FG may in general be reliable and reproducible. As expected, however, registry data did not have the granularity to differentiate distinct courses of AD present among the trial participants. Another notable insight was that in only one of the 12 participants with ICD code F06.7 the mild cognitive disorder was attributable to AD. Researchers often consider mild cognitive impairment as a transitory phase from normal cognition to AD if no other obvious reasons for the cognitive deficits can be identified. We note that, while considered a proxy endpoint, F06.7 subsumes a broader disease spectrum than the mild cognitive impairment as defined in the National Institute on Aging – Alzheimer’s Association guidelines (30). In fact, our results propose that Finnish clinicians use this code to document the presence of cognitive impairment as part of medical conditions other than AD, reflected by eight different probable underlying conditions among the 12 individuals in the MCD subgroup. Our findings thus clearly advise against using F06.7 as a code with the aim to identify early or pre-symptomatic AD patients from a biobank cohort.

For participants who completed the cognitive assessments via in-person, computerized and telephone-based testing, we observed a high correlation between MMSE and TELE. This is consistent with a previous study in Finnish patients with AD and healthy controls (17). In comparison to MMSE and TELE, the CERAD-NB word list cut-off for immediate and delayed recall identified higher numbers of cognitively impaired individuals in both subgroups, emphasizing that studies like ours benefit from including more specific memory measures rather than simply relying on short screening instruments. Moderate to strong correlations between in-person (CERAD-NB) and computerized (cCOG) measures of immediate and delayed recall are in line with earlier studies comparing computerized/web-based versus in-person neuropsychological tests (14, 31–34). We also observed moderate to strong correlations between telephone-administered and in-person episodic memory measures. To our knowledge, our study is the first to use three learning trials of the 10-word list learning task included in the TICS-m. Thus, our telephone-based episodic memory measures closely corresponded to in-person word-list episodic memory measures. The lowest correlations between episodic memory measures were between TICS-m and cCOG word list measures: this could result from the fact these measures are based solely on verbal versus visual stimuli, respectively, whereas CERAD-NB word list includes both, visual and verbal stimuli. Based on the substantial inter-platform variability we observed in our study, future studies should be advised to assess episodic memory with more than a single test.

We observed significant associations between cCOG and in-person measures also for other cognitive domains. Processing speed, as assessed by TMT-A, showed a moderate association between in-person and cCOG assessment after exclusion of outliers. For TMT-B, measuring set-shifting, one of the core executive functions, showed a strong correlation between cCOG and in-person testing, which remained after accounting for processing speed and exclusion of outliers from TMT-A. These results are consistent with earlier findings (14). However, given that a sizable fraction of participants was unable to complete the task, computer-based administration of TMT-A/B in an unfamiliar environment on site may be considered as too difficult for individuals with manifest AD or MCD. Notably though, we observed a strong association between in-person and telephone administered semantic fluency. Due to the fact that the 1-minute animal naming was identical in both formats, this measure represented the strongest correlation among the measures of specific cognitive abilities. We are not aware of any other studies that have examined the association between in-person and telephone administered semantic fluency.

Despite the small size of our study, plasma levels of GFAP and pTau-181 were significantly different between the AD and MCD groups, which further supports the potential of these measures as diagnostic biomarkers for AD (35). Conversely, no such differentiation was seen for Aβ42/40 ratio and NfL. Our results thus back recent findings that among the four plasma parameters pTau-181 might be the strongest differentiator of AD from non-AD patients (36), which will need to be further validated at larger sample sizes. Larger studies will also be needed to thoroughly compare blood-based AD biomarker levels to cognitive performance. Notably, however, already in our current study we observed negative associations of TELE total score with GFAP, NfL, and pTau-181 levels, yet not with Aβ42/40 ratio. According to the expectations, there was a trend towards more *APOE* ε4-carriers in the AD group than in the MCD group, with the prevalence of ε4-carriers in the MCD group closely matching to the prevalence of over 30% in the Finnish population (21).

Our study has several limitations: First, we invited participants from just a single of the nine Finnish biobanks contributing to FG and focused recruitment on a relatively uniform cohort in geographic proximity to the single study center. Follow-up studies that aim for a larger and more diverse study population will certainly face additional complexities, not the least with regards to harmonizing re-contacting, consenting, and contracting activities between multiple parties. It can be expected that the establishment of a centralized coordinating unit, such as the Finnish Biobank Cooperative FinBB, will facilitate the navigation through complex administrative processes that have not yet been customized for clinical recall activities in Finland and in our case required an extended period of planning. Second, according to FG records all individuals in our target population had already been diagnosed with either AD or MCD as a reflection of cognitive deficiencies. One of the greatest opportunities of biobank studies is to facilitate contact to individuals early in their disease or at risk, but not yet manifest disease, for instance because they carry genetic variants that without intervention reduce lifespan (8,37,38). In fact, enrolling sufficient “trial-ready” minimally symptomatic AD patients into interventional trials often takes several years and may contribute substantially to the costs of clinical programs (39).

While FG participants are in principle consented for return of their results, studies in pre-and early-symptomatic individuals will need to carefully walk the fine line between providing useful information to improve a participant’s health or lifestyle and disclosing potentially unwanted and uncertain research information (40). The correspondence between measures of cognitive function across different formats used in our study was generally good, we observed substantial variability of cognitive performance among the small number of individuals recalled for our study. This at least in part reflects that a few study participants were substantially further progressed in their disease than others. Further stratification based on available FG data may make future cohorts more homogenous. Also, cognitive function information acquired by phone in general tended to correlate better to in-person tests than information obtained via cCOG in our cohort; this may be due to our sample of individuals with AD having more problems with self-guided computerized tool versus using the phone. Finally, we note that with its small sample size our pilot study was not designed to detect robust statistical differences between measures, so results should be evaluated with caution.

In summary, our study introduces a recall framework to launch clinical studies from FG and the Finnish biobanks. While laborious to operationalize, the opportunity to recontact FG participants based on distinct genetic or phenotypic parameters opens the door for a broad spectrum of scientific activities, ranging from the validation of genetic findings in targeted follow-up experiments to delivering new and emerging precision treatments to individuals with the highest medical needs.

## Data Availability

All data produced are available in FinnGen sandbox, a safe controlled environment accessible by authorized researchers in FinnGen research community.

## ACKNOWLEDGEMENTS

We are grateful to the participants of FinnGen and this current study for volunteering their data to research. We further thank Mary Pat Reeve for FinnGen Sandbox look-ups and Huei-Yi Shen for help with contracting. The FinnGen project is funded by two grants from Business Finland (HUS 4685/31/2016 and UH 4386/31/2016) and the following industry partners: AbbVie, AstraZeneca UK, Biogen, Bristol Myers Squibb (and Celgene Corporation & Celgene International II), Genentech, Merck Sharp & Dohme LLC, a subsidiary of Merck & Co., Inc., Rahway, NJ, USA, Pfizer, GlaxoSmithKline Intellectual Property Development, Sanofi US Services, Maze Therapeutics, Janssen Biotech, Novartis, and Boehringer Ingelheim. The following biobanks are acknowledged for delivering samples to FinnGen: Auria Biobank (https://www.auria.fi/biopankki/), THL Biobank (https://www.thl.fi/ biobank), Helsinki Biobank (https://www.helsinginbiopankki.fi), Biobank Borealis of Northern Finland (https://www.ppshp.fi/Tutkimus-ja-opetus/Biopankki/Pages/Biobank-Borealis-briefly-in-English.aspx), Finnish Clinical Biobank Tampere (https://www.tays.fi/en-US/Research_and_development/Finnish_Clinical_Biobank_Tampere), Biobank of Eastern Finland (https://www.ita-suomenbiopankki.fi/en), Central Finland Biobank (https://www.ksshp.fi/fi-FI/Potilaalle/Biopankki), Finnish Red Cross Blood Service Biobank (www.veripalvelu.fi/verenluovutus/biopankkitoiminta) and Terveystalo Biobank (https://www.terveystalo.com/fi/Yritystietoa/Terveystalo-Biopankki/Biopankki/). All Finnish biobanks are members of the BBMRI.fi infrastructure (https://www.bbmri.fi). The FINBB (https://finbb.fi/) is the coordinator of BBMRI-ERIC operations in Finland. The Finnish biobank data can be accessed through the Fingenious services (https://site.fingenious.fi/en/) managed by FINBB. We acknowledge the BBMRI.fi for creating the original biobank map used in Fig. 1a and providing us their permission to use it. Further funding to EV came from the Academy of Finland grants 314639 and 320109.

The FinnGen study is approved by Finnish Institute for Health and Welfare (permit numbers: THL/2031/6.02.00/2017, THL/1101/5.05.00/2017, THL/341/6.02.00/2018, THL/2222/6.02.00/2018, THL/283/6.02.00/2019, THL/1721/5.05.00/2019 and THL/1524/5.05.00/2020), Digital and population data service agency (permit numbers: VRK43431/2017-3, VRK/6909/2018-3, VRK/4415/2019-3), the Social Insurance Institution (permit numbers: KELA 58/522/2017, KELA 131/522/2018, KELA 70/522/2019, KELA 98/522/2019, KELA 134/522/2019, KELA 138/522/2019, KELA 2/522/2020, KELA 16/522/2020), Findata permit numbers THL/2364/14.02/2020, THL/4055/14.06.00/2020,,THL/3433/14.06.00/2020, THL/4432/14.06/2020, THL/5189/14.06/2020, THL/5894/14.06.00/2020, THL/6619/14.06.00/2020, THL/209/14.06.00/2021, THL/688/14.06.00/2021, THL/1284/14.06.00/2021, THL/1965/14.06.00/2021, THL/5546/14.02.00/2020, THL/2658/14.06.00/2021, THL/4235/14.06.00/202, Statistics Finland (permit numbers: TK-53-1041-17 and TK/143/07.03.00/2020 (earlier TK-53-90-20) TK/1735/07.03.00/2021, TK/3112/07.03.00/2021) and Finnish Registry for Kidney Diseases permission/extract from the meeting minutes on 4th July 2019. The Biobank Access Decisions for FinnGen samples and data utilized in FinnGen Data Freeze 9 include: THL Biobank BB2017_55, BB2017_111, BB2018_19, BB_2018_34, BB_2018_67, BB2018_71, BB2019_7, BB2019_8, BB2019_26, BB2020_1, Finnish Red Cross Blood Service Biobank 7.12.2017, Helsinki Biobank HUS/359/2017, HUS/248/2020, Auria Biobank AB17-5154 and amendment #1 (August 17 2020), AB20-5926 and amendment #1 (April 23 2020) and it’s modification (Sep 22 2021), Biobank Borealis of Northern Finland_2017_1013, Biobank of Eastern Finland 1186/2018 and amendment 22 § /2020, Finnish Clinical Biobank Tampere MH0004 and amendments (21.02.2020 & 06.10.2020), Central Finland Biobank 1-2017, and Terveystalo Biobank STB 2018001 and amendment 25th Aug.

## AUTHOR CONTRIBUTIONS

V.J., E.V. and H.R. conceptualized and oversaw the study; V.J., C.S. and E.V. conducted statistical analyses; V.J., J.K., M.H., A.-K.P., A.M., H.K., T.L., V.-M.K., T.I.P., R.K., M.H., S.-K.H., S.K., T.K., M.U., obtained or contributed data or facilitated interactions at participating institutions; M.P. and A.P. consulted on the study protocol and enabled governance interactions; V.J., C.S., E.V. and H.R. wrote and edited the study protocol and manuscript

## CONFLICTS

H.R. is a full-time employee of Biogen. A.P. is a member of the Pfizer Genetics Scientific Advisory Panel.

## FinnGen Collaborators

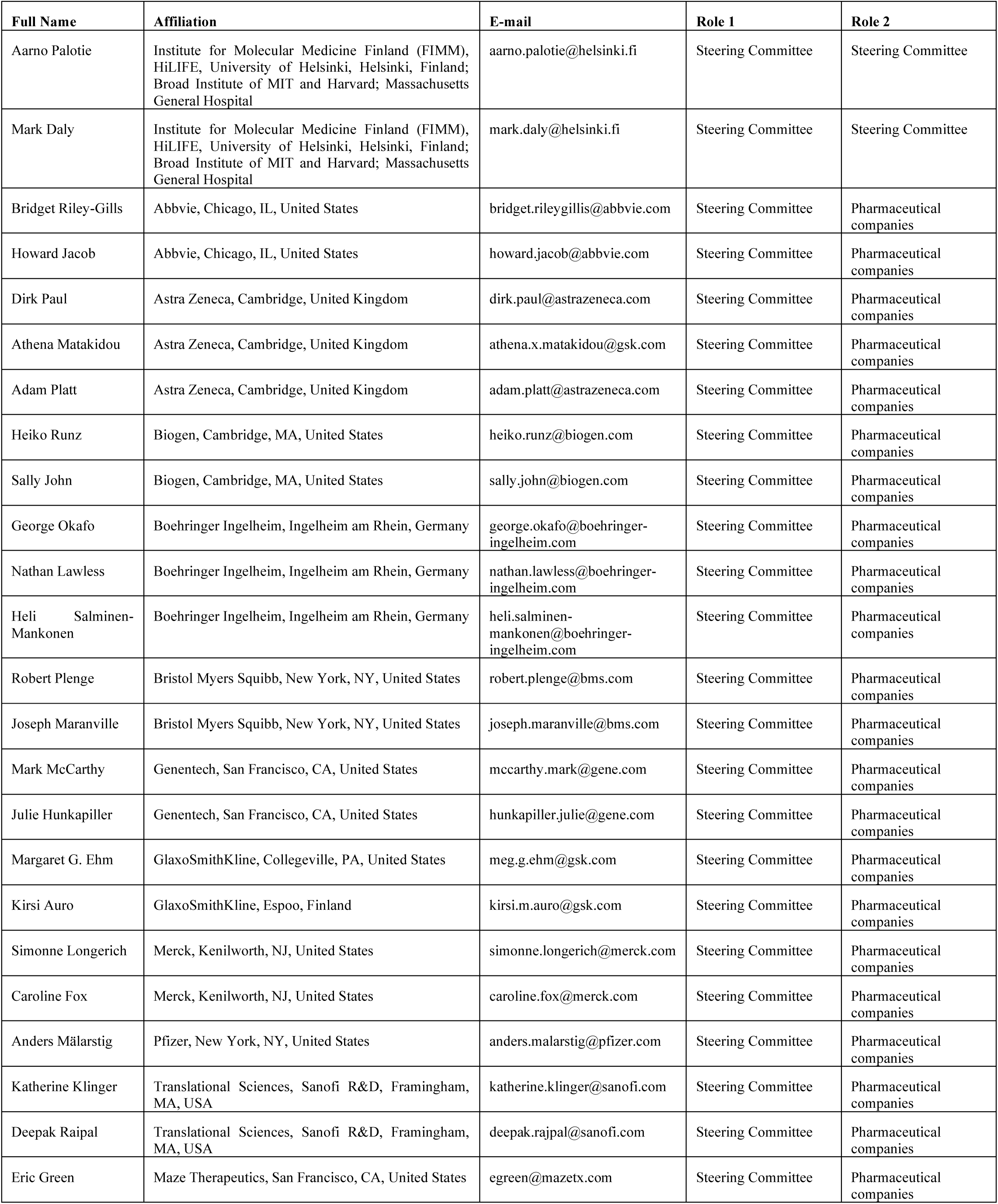
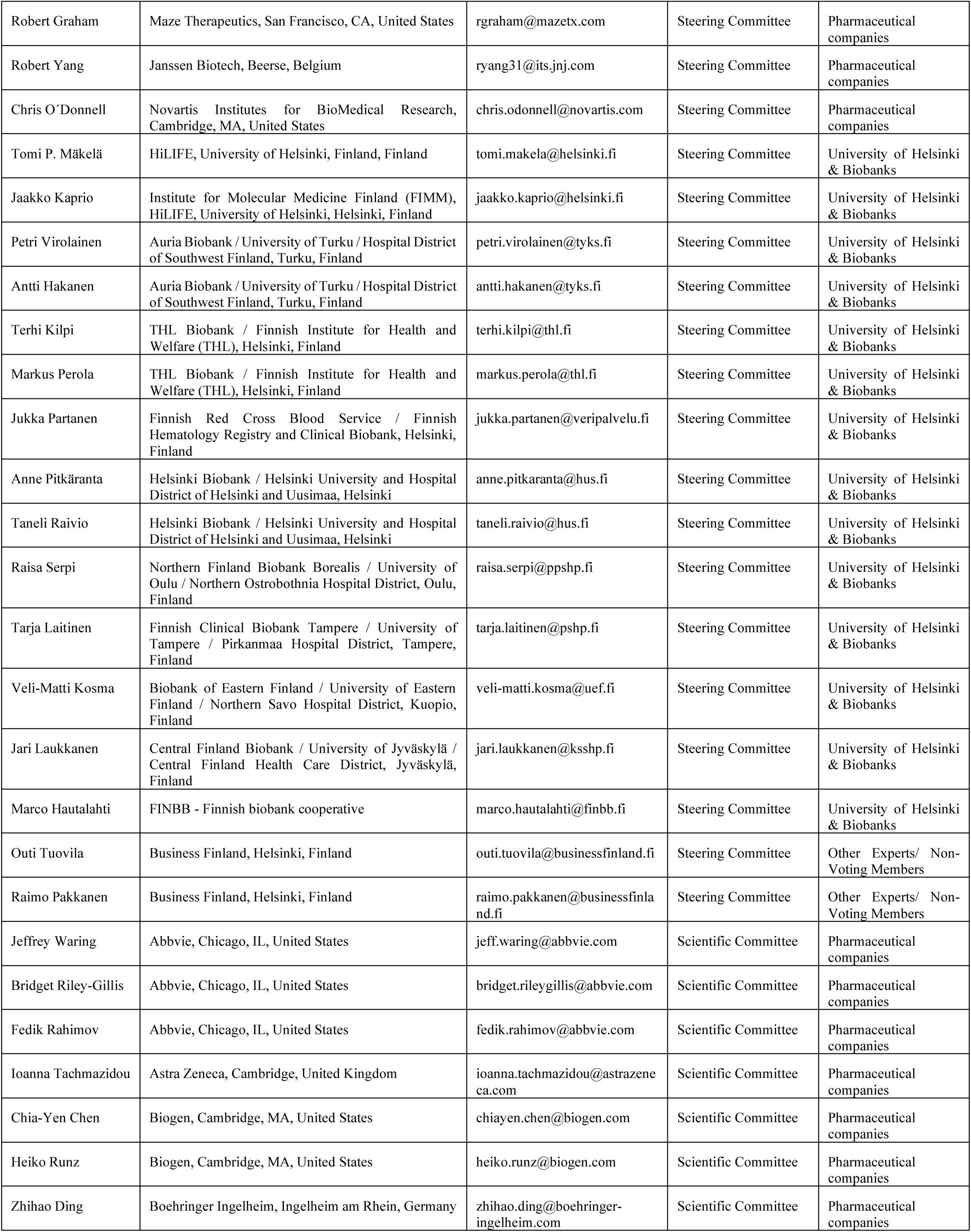
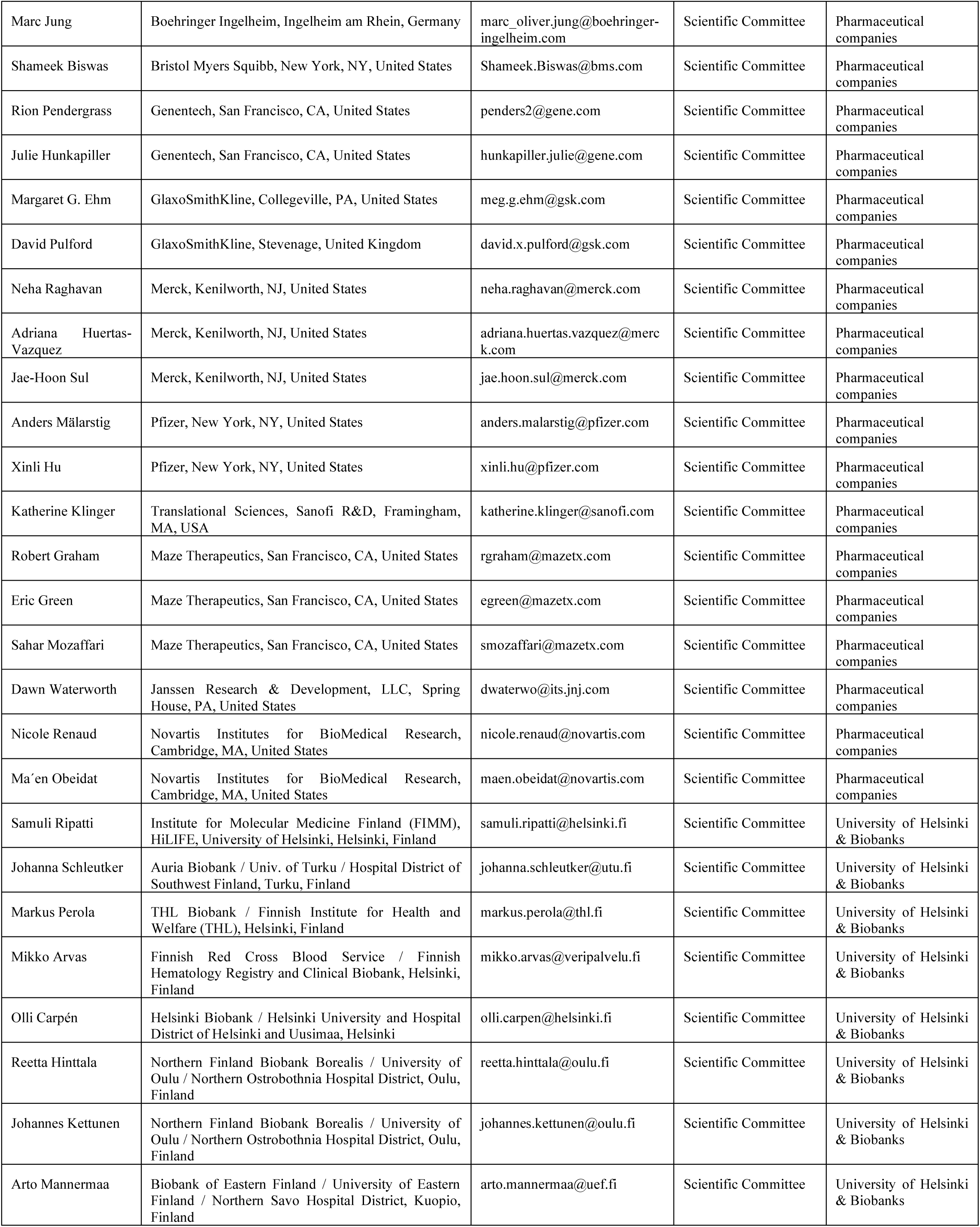
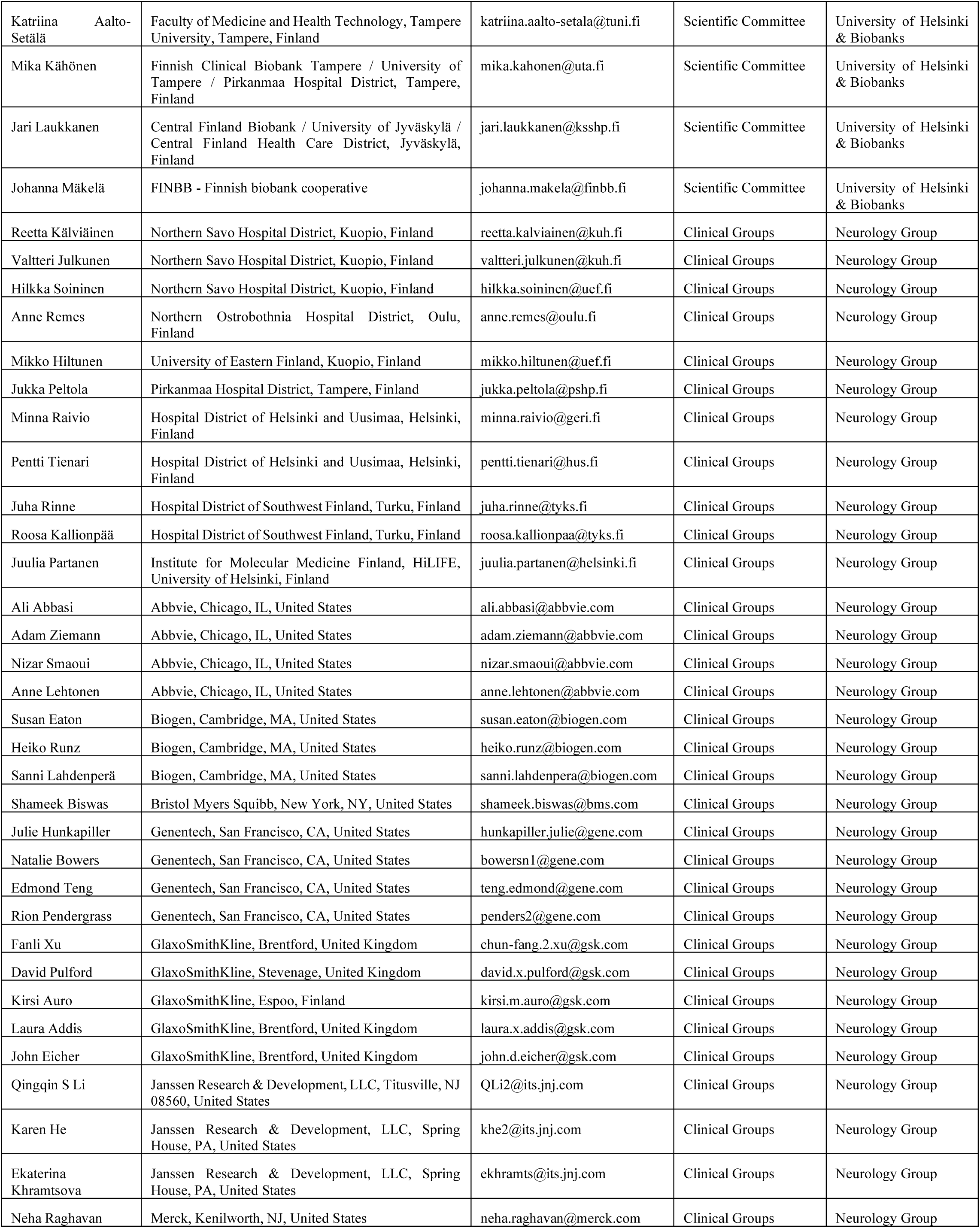
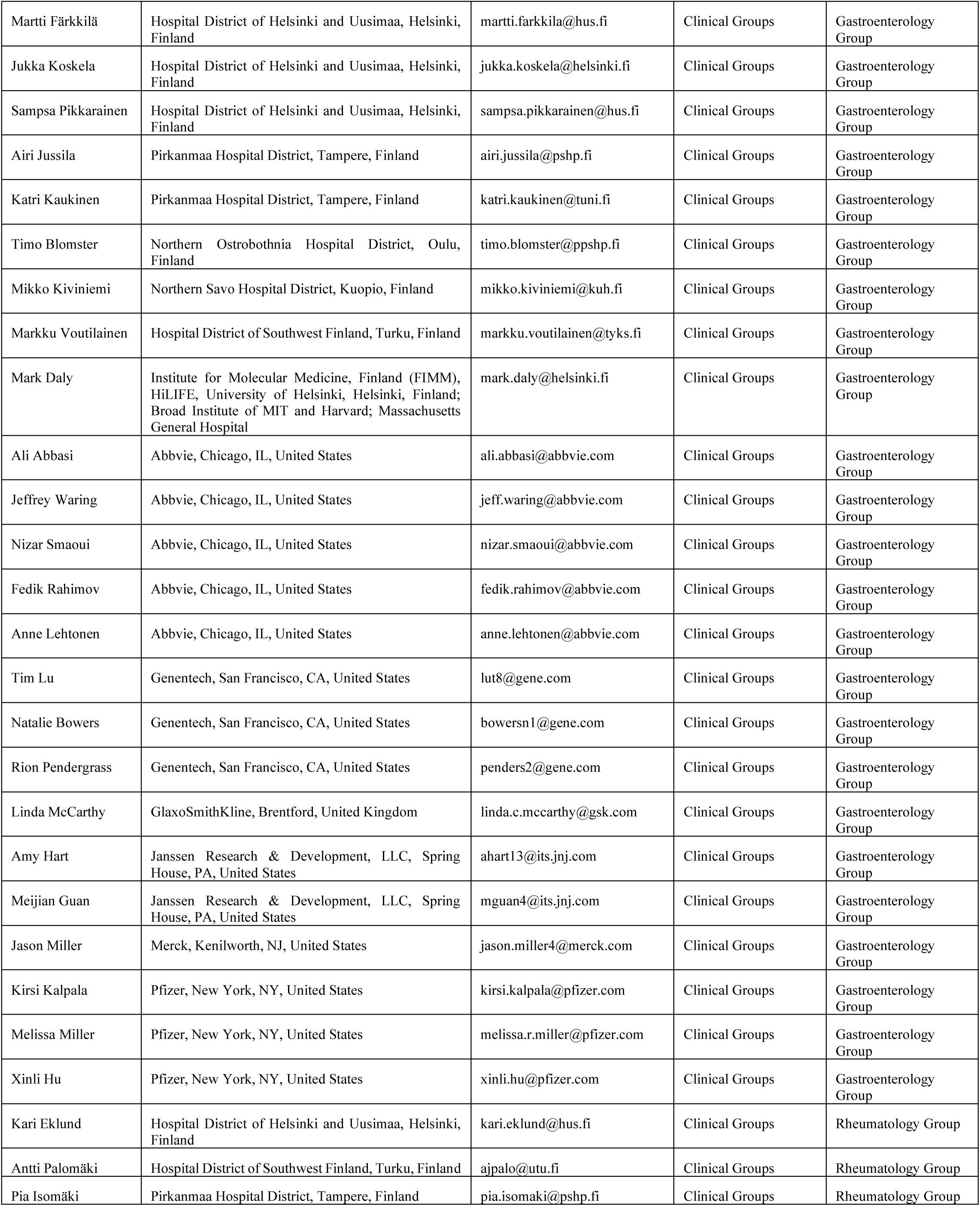
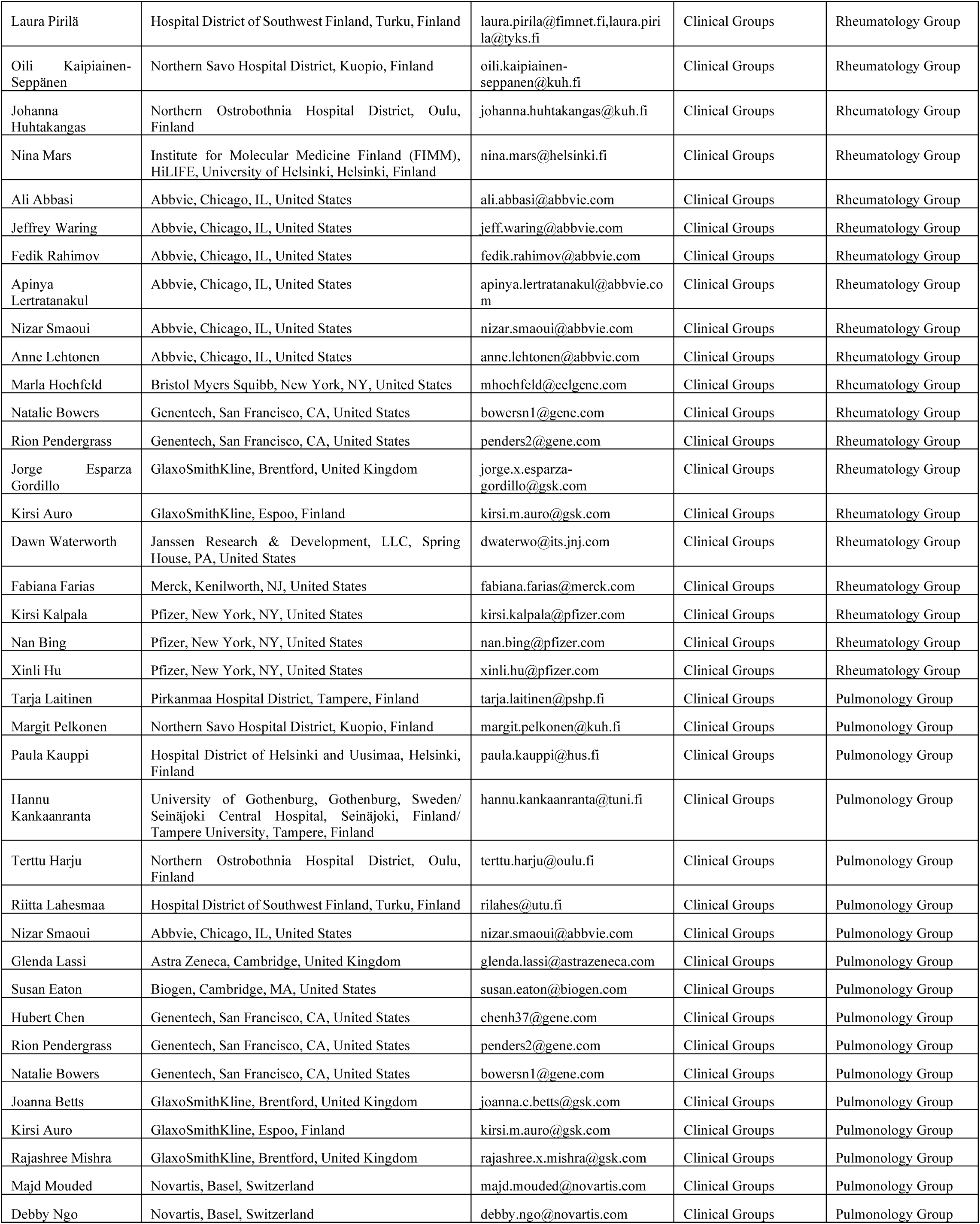
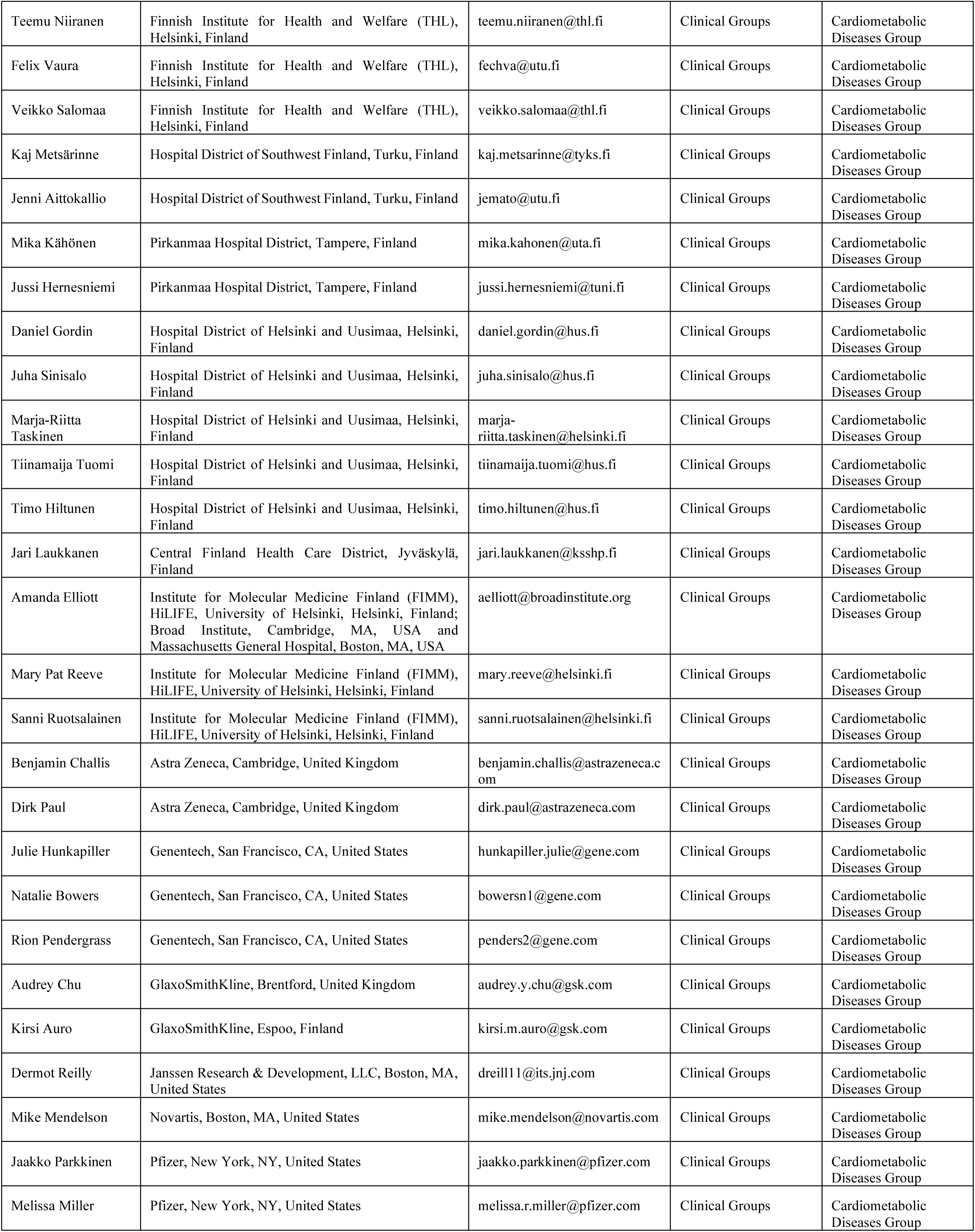
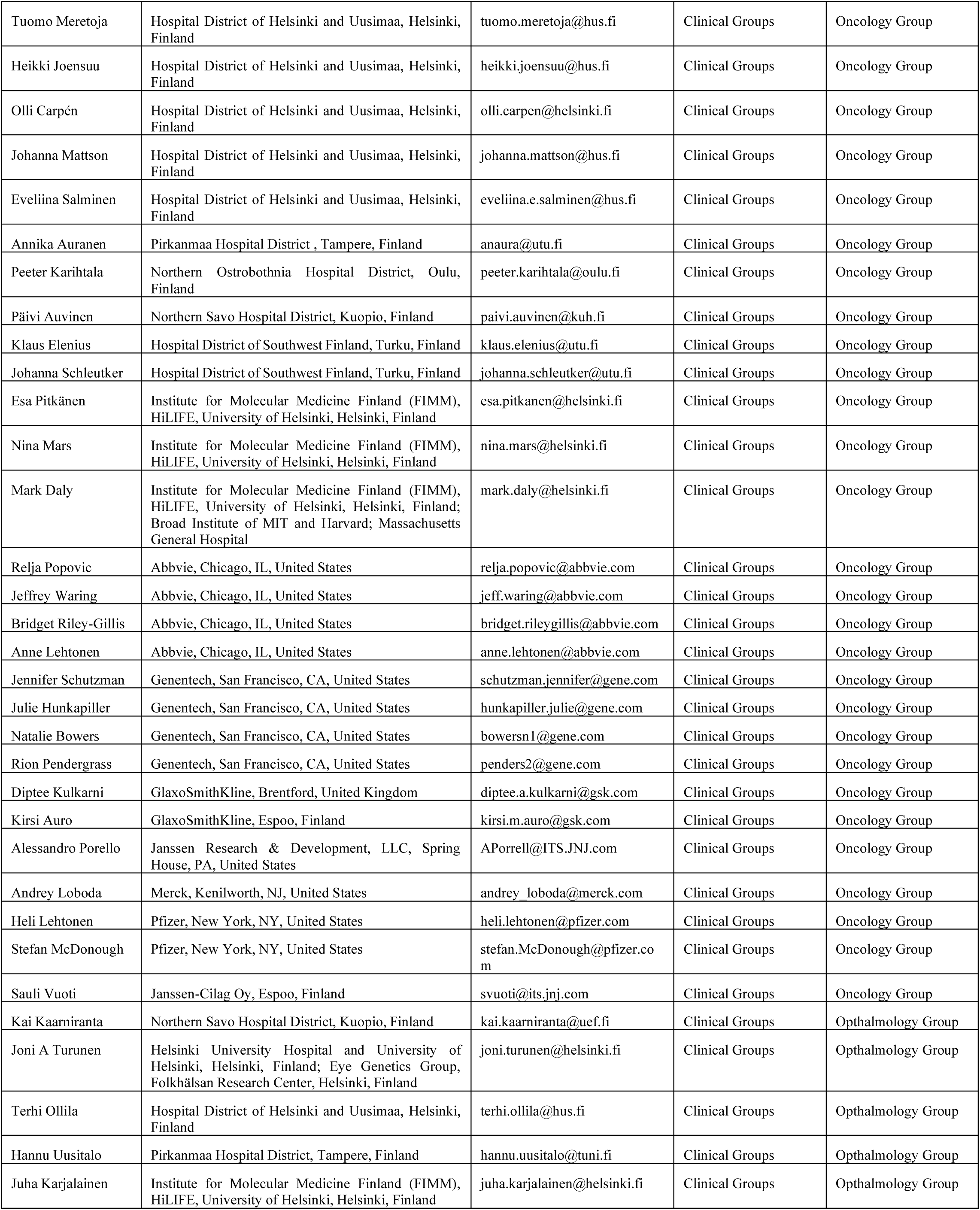
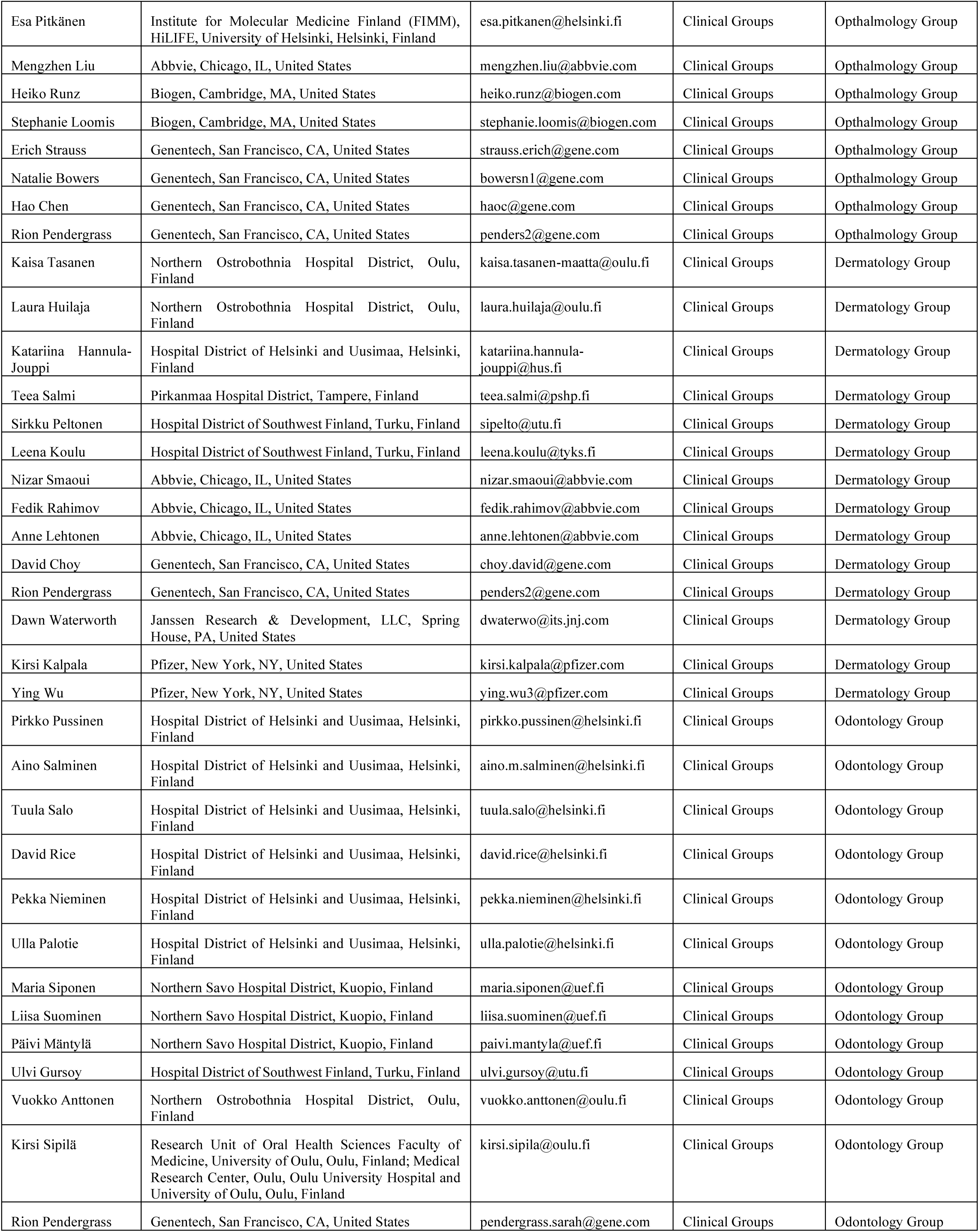
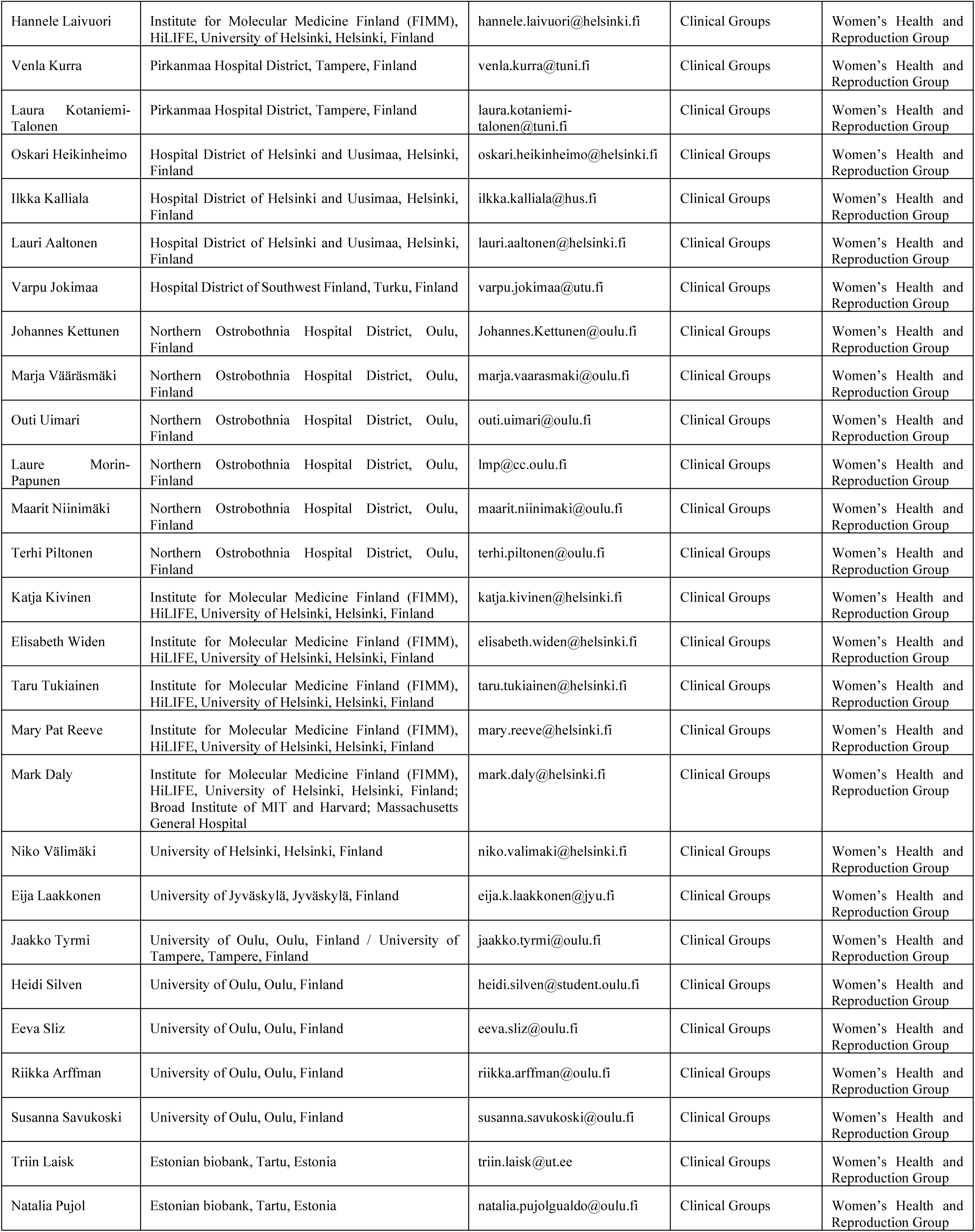
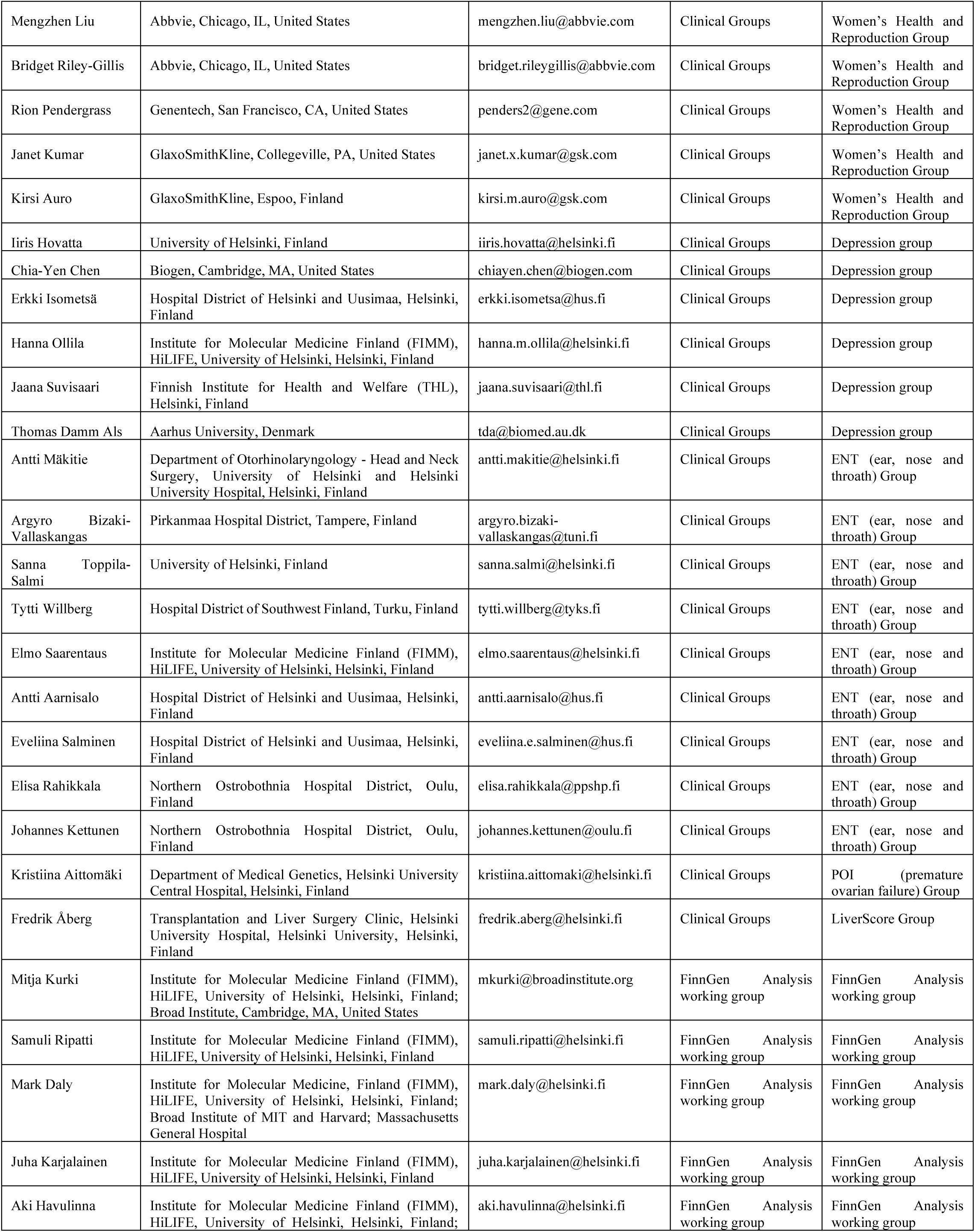
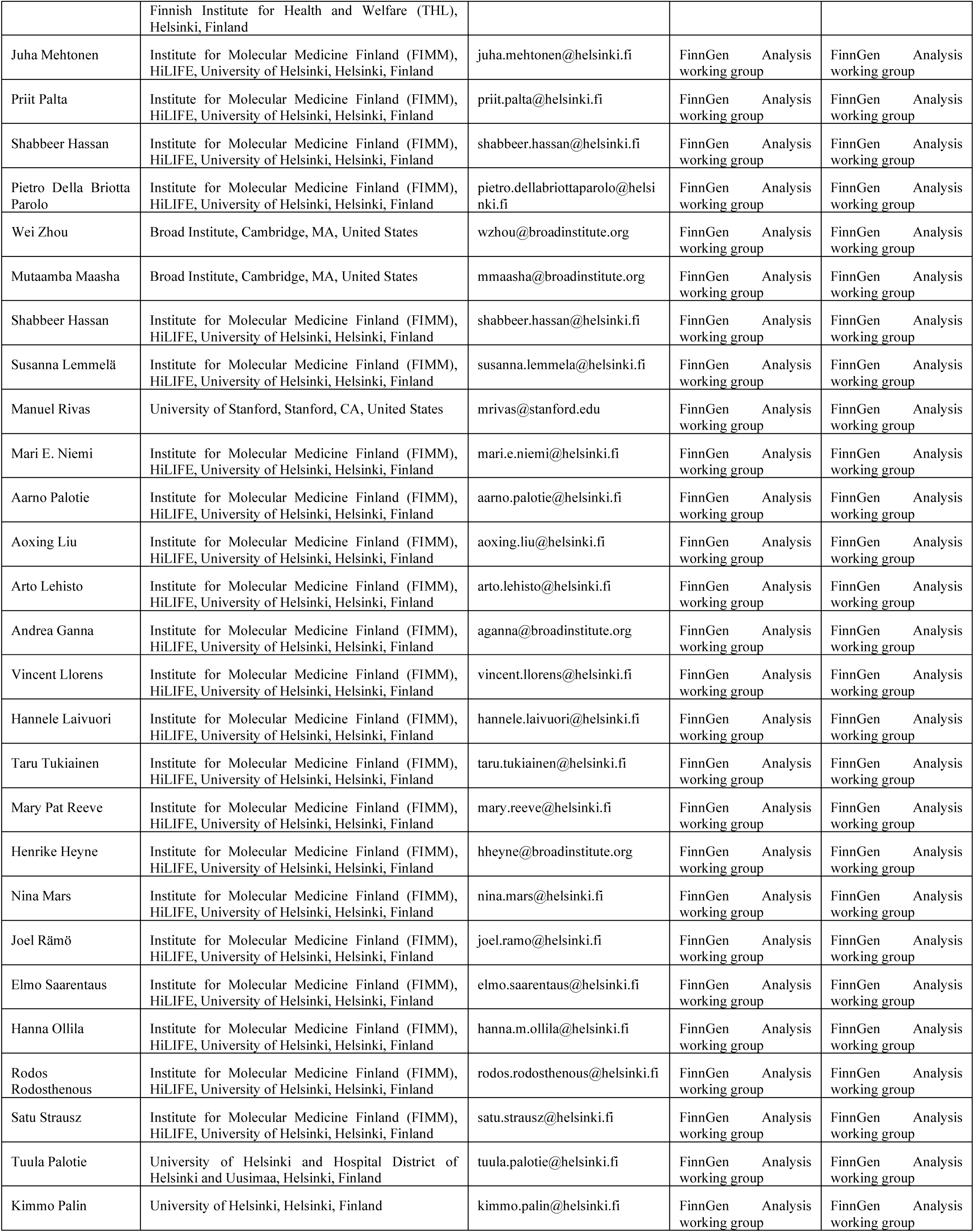
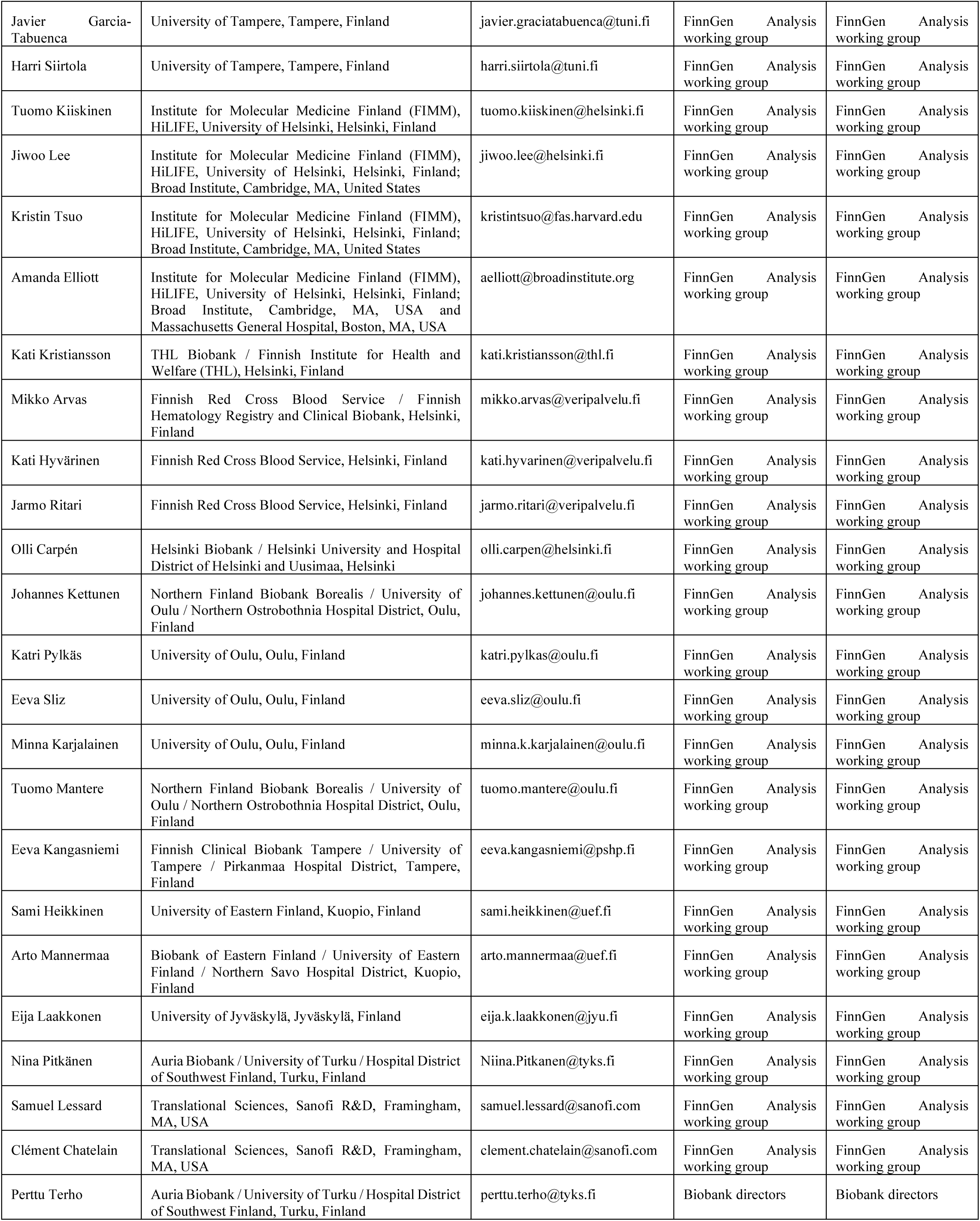
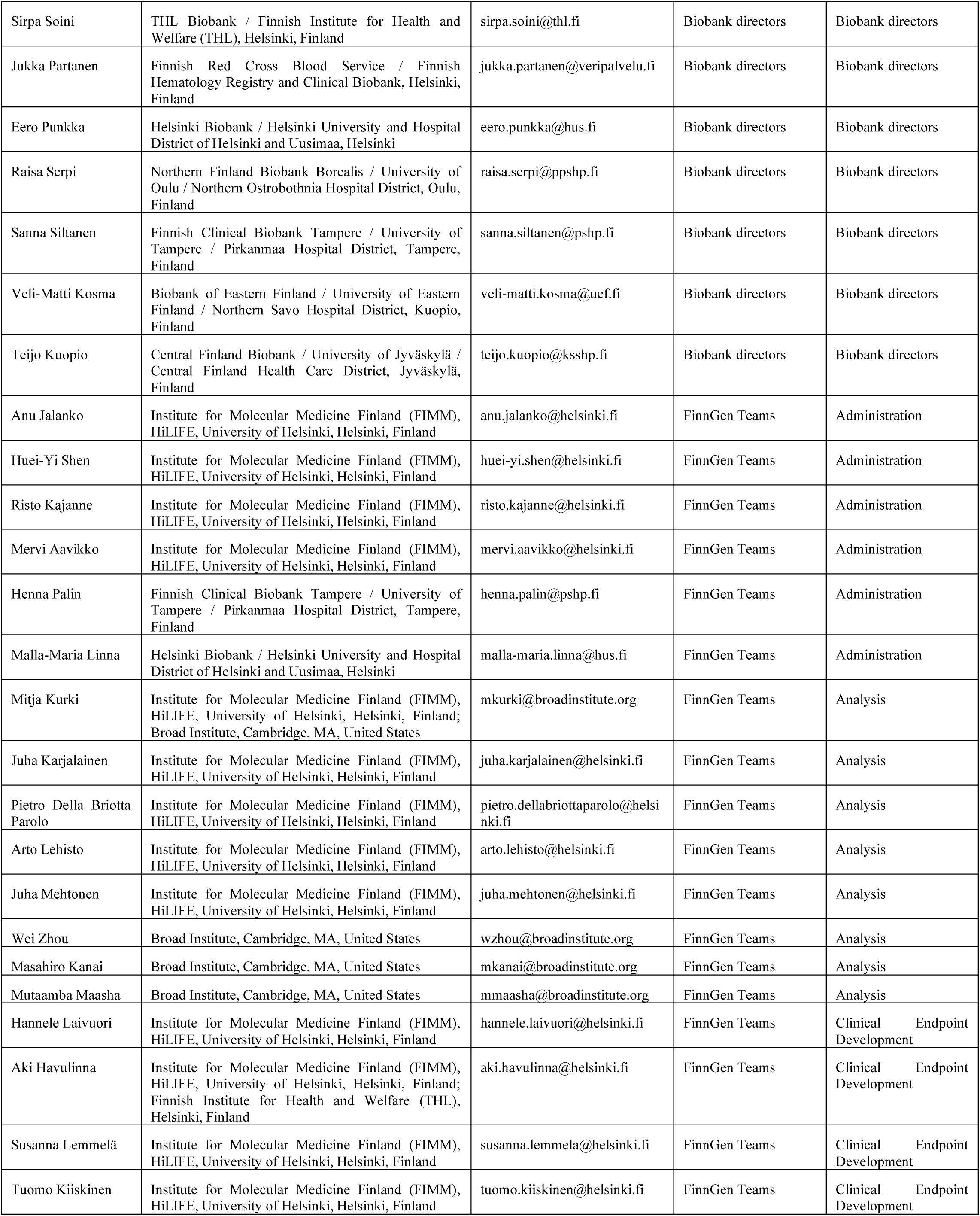
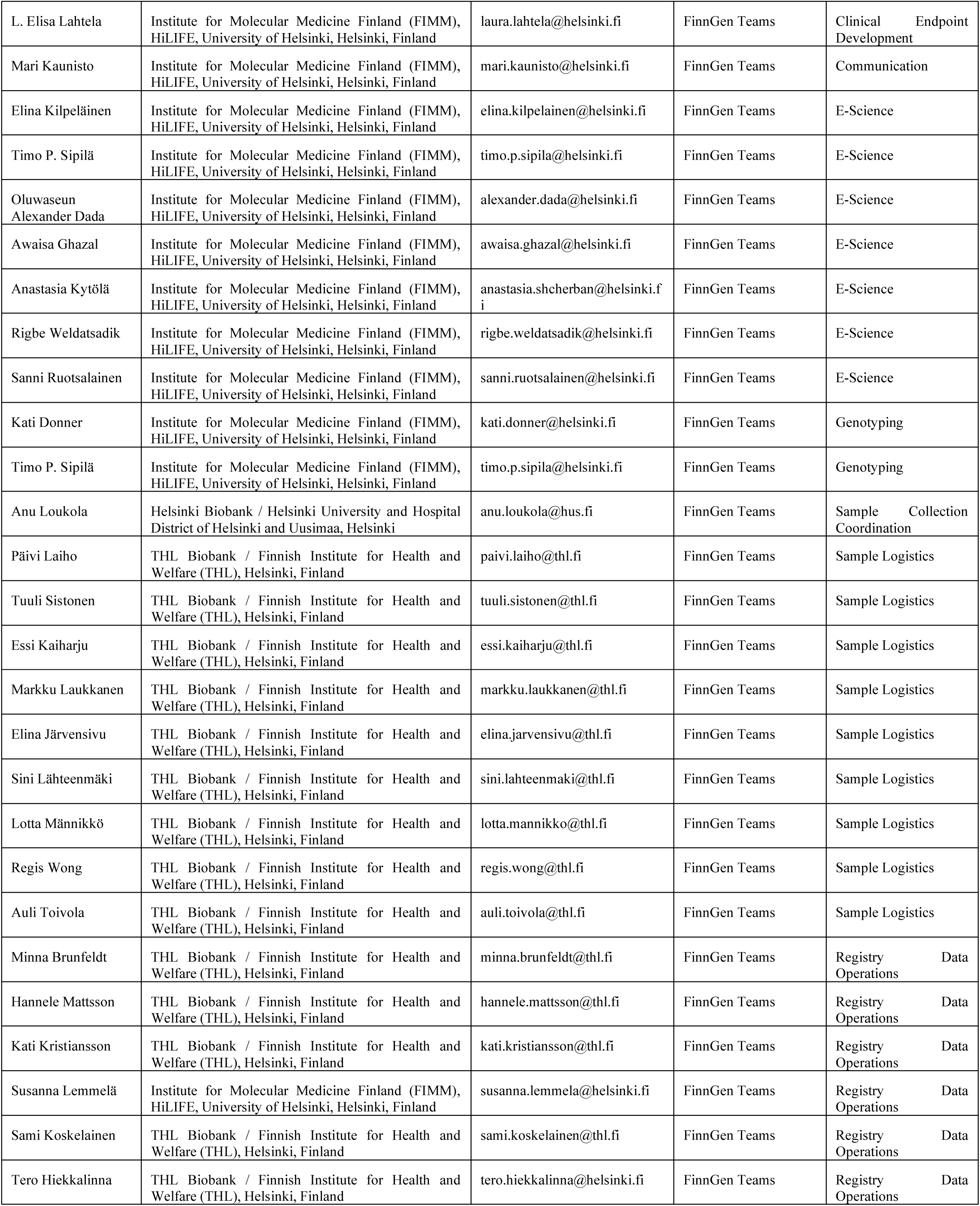
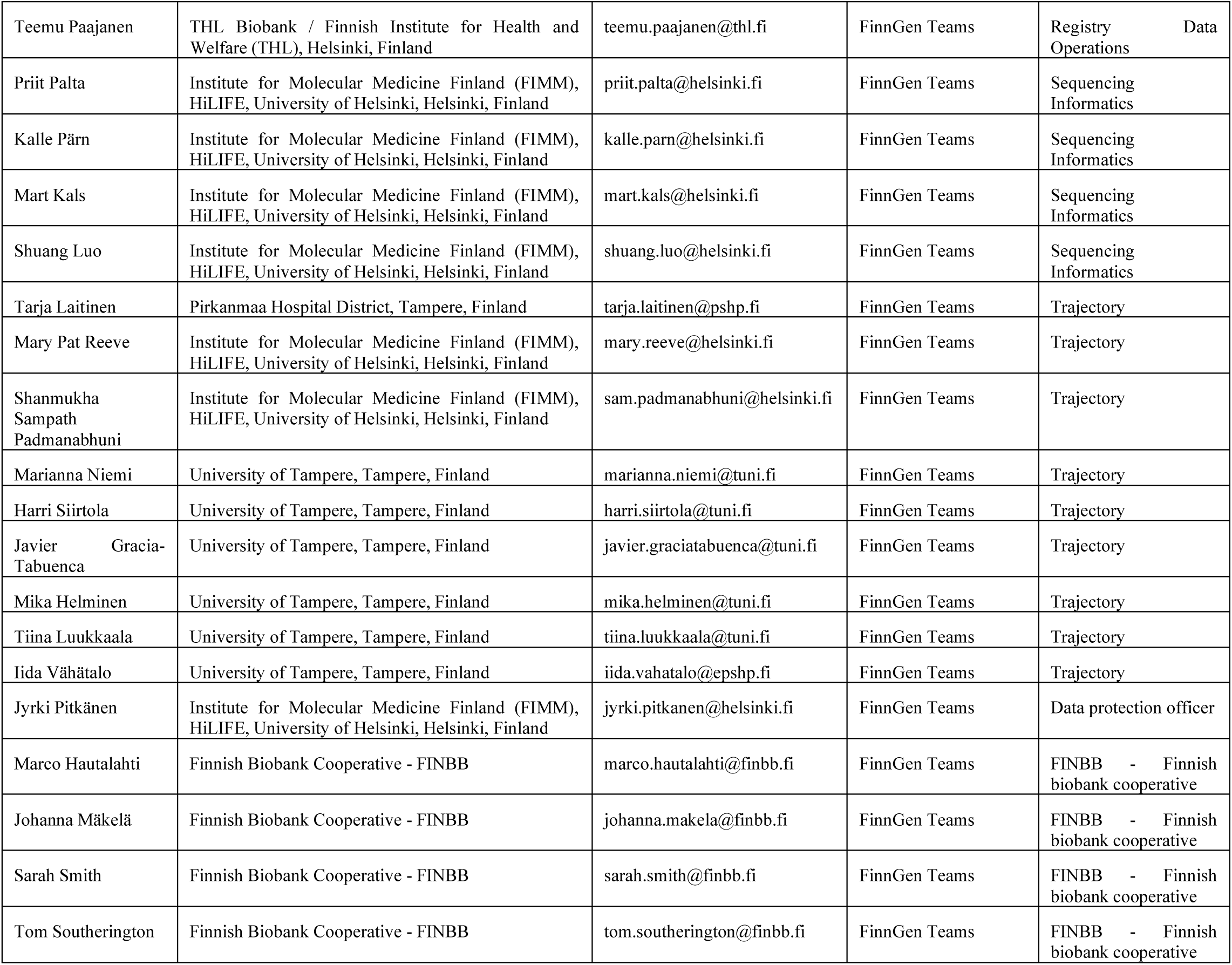

## Notes

### Funding Statement

This study was funded by the FinnGen project. The FinnGen project is funded by two grants from Business Finland (HUS 4685/31/2016 and UH 4386/31/2016) and the following industry partners: AbbVie, AstraZeneca UK, Biogen, Bristol Myers Squibb (and Celgene Corporation & Celgene International II), Genentech, Merck Sharp & Dohme LLC, a subsidiary of Merck & Co., Inc., Rahway, NJ, USA, Pfizer, GlaxoSmithKline Intellectual Property Development, Sanofi US Services, Maze Therapeutics, Janssen Biotech, Novartis, and Boehringer Ingelheim. Further funding to EV came from the Academy of Finland grants 314639 and 320109.

### Author Declarations

Ethics committee of HUS gave ethical approval for this work. Additionally, research permission was granted by from the Biobank of Eastern Finland and Kuopio University Hospital.

